# Characterization of Body Composition Dynamics Throughout Treatment in Patients with Early-Stage Breast Cancer

**DOI:** 10.1101/2025.07.30.25332446

**Authors:** Lauren E. Rentz, McKinzey K. Dierkes, Beth Vettiyil, Ida Holásková, Emidio E. Pistilli

**Author notes:** Correspondence; Tel.: +1−304−293−0291.

## Abstract

This study characterized computed tomography (CT)-derived body composition and tissue morphology in females with non-metastatic breast cancer (BC), both cross-sectionally (n = 56) and longitudinally (n = 38), with comparative evaluations against healthy controls and cachexia-prone females with lung cancer. Despite relative weight stability, BC patients demonstrated significant declines in skeletal muscle quality and increases in subcutaneous adipose attenuation. Morphologic changes occurred even in the absence of pronounced muscle loss, namely through reductions in lean tissue masked by concurrent increases in intra-compartmental adipose. Longitudinal interactions suggest divergent phenotypes such that non-cachectic patients demonstrated subtle, though measurable, remodeling of stable muscle quantity, whereas wasting of both muscle and adipose quantities and remodeling of externally deposited adipose was characteristic of cachexia. Findings highlight the discordance between weight loss and underlying tissue morphology and suggest that conventional cachexia criteria may under-detect clinically relevant remodeling in early-stage BC. CT-based assessments may improve phenotyping and better inform supportive care strategies across cancer populations.

## INTRODUCTION

A disease that will ultimately impact 1 in 8 women, breast cancer (BC) represents the malignancy with the largest population of disease-free survivors, with a surmountable 274,780 patients becoming survivors in 2025 alone.^1^ The 86% 10-year survival rate is attributable to decades of research regarding the screening, diagnostics, and treatment of BC, and reflects a major achievement in combating this disease. Albeit, while mortality rates have decreased by more than 40% in the last few decades, incidence rates have continued to rise.^1,2^ As such, the widespread burden of long-lasting disease implications continues to hinder survivors’ quality of life for decades following cessation of treatment, and thus, affecting more women now than ever before.

Clinical presentation of BC differs from many cancer types, particularly in the progression of disease and treatment options, given most cases are caught prior to metastasis.^2^ More specifically, early-stage BC patients share many symptoms with other cancer types, such as malaise, a loss of appetite and fatigue, though differ in that body weight is often stable during active treatment. Interestingly, nearly all BC patients report fatigue to some degree;^3^ however, considering early-stage treatment almost always includes surgical resection, tissue wasting is thought to be uncommon in this population. Rather, cancer-cachexia, most commonly defined as secondary weight loss that exceeds five percent of a patients’ initial body weight,^4^ is a greater concern for more advanced stages of BC, including metastatic disease.^5^

Notably, symptomatic fatigue has been reported to have the most substantial impact on patient lives, affecting nearly all individuals with BC throughout the management of the disease.^6,7^ For some, this fatigue persists well beyond a decade after treatment cessation.^3^ As such, understanding the mechanisms and manifestations of cancer-related fatigue is critical for advancing therapeutics, specifically those capable of optimizing disease prognosis without sacrificing the quality of life. Previously, our laboratory has characterized a phenotype of BC-induced skeletal muscle fatigue in the absence of muscle wasting among patients with early-stage BC, which exists independent of tumor subtype or the use of directed therapies (i.e., chemotherapy, radiation, etc.). Across numerous animal models, this phenotype is characterized by greater fatiguability of skeletal muscle, despite no difference in muscle weights compared to healthy mice. Further, this fatiguability presents with genomic and proteomic profiles indicative of metabolic and mitochondrial dysfunction; these multi-omic profiles of dysfunction have been recapitulated in skeletal muscle from early-stage BC patients, and has been responsive to drug therapy in the pre-clinical setting.^8–10^

We have also contributed to the vast body of literature surrounding the characterization of body composition and changes to body mass among early-stage BC patients. We previously demonstrated widespread weight stability among thousands of patients, in addition to bioelectrical impedance (BIA)-estimated body composition that was comparable to female controls without BC.^11^ Our prior work, along with others, suggests relative stability in body mass in early-stage BC patients.^12–14^ However, variability exists across studies, with some groups reporting weight loss^15^ and some weight gain^16^ among similar patient samples.

Segmentation of computed tomography (CT) has been used extensively in clinical populations for estimation of body composition, and is considered a gold standard approach.^17,18^ Despite this, CT imaging is rarely ordered in patients with early-stage BC, making data availability scarce for this patient cohort. As such, retrospective image analysis of non-metastatic BC patients may pose challenges through limited data availability and a lack of consistency.^19^ Retrospectively obtained samples from electronic medical records may also be susceptible to selection bias, which likely over-represents patients with more advanced disease characteristics. Among studies utilizing CT-derived estimates, many entail scans ordered during follow-up visits, where CT imaging is most prevalent with respect to standard of care procedures. Follow-up CTs are often used to screen for disease reoccurrence and metastasis; so while more common, these scans tend to represent BC survivors, rather than active, early-stage disease. As such, accurate quantification of body compositional changes in non-metastatic BC patients remains scarce, primarily due to limited data availability.

Despite an abundance of evidence that cancer cachexia occurs secondary to most major cancers,^20^ this finding has not been consistently shown to extend to early-stage BC, despite the abundance of evidence suggesting fatigue and functional declines.^3,21^ Therefore, we seek to further investigate changes in body composition occurring in the early stages of BC by utilizing numerous between-patient and within-patient evaluations to characterize body composition and tissue morphology. We hypothesize that changes to body composition, including skeletal muscle composition, will occur among female patients diagnosed with non-metastatic BC throughout neoadjuvant treatment. Specifically, pre-treatment characterization of body tissues among non-metastatic BC patients is expected to be comparable in quantity to that of healthy females. Further, we expect some variability in muscle quality, which may more-closely mirror females diagnosed with aggressive malignancies associated with poorer outcomes. Finally, early-stage BC patients are expected to better retain tissue quantities during treatment and undergo less dramatic changes to body composition than females with other malignancies. In this study, we seek to provide multi-faceted, longitudinal CT-derived characterizations of body composition in BC patients and comparatively in other female cohorts, as well as highlight the corresponding tissue morphology with clinically derived stratifications for cachexia.

## METHODS

### Patients

All data and images utilized herein were derived from open-access datasets made available by The Cancer Imaging Archive (TCIA).^22^

The primary study sample was drawn from the *ACRIN-FLT-Breast dataset*,^23^ which originates from the multi-center ACRIN-6688 clinical trial, which sought to compare tumor pathologic response with PET imaged FLT uptake.^24,25^ This cohort was used to evaluate non-metastatic BC both cross-sectionally and longitudinally. This patient cohort represents females with a confirmed diagnosis of stage IIA-IIIC BC scheduled to receive NAC prior to surgical tumor resection. All patients were determined to have normal organ function at enrollment, were devoid of uncontrolled intercurrent illness, and had no personal history of malignancy or cancer treatment. Patients were imaged with CT from skull-to-thigh prior to initiating personalized NAC (*pre-NAC*), and again following NAC completion (*post-NAC*; within 3 weeks of surgical resection).^24^ Patients with a *pre-NAC* scan free of artifact and with measurements of body height and weight from the day of the scan were included in our cross-sectional comparisons (n = 56). Further, patients imaged at both *pre-NAC* and *post-NAC* timepoints obtained at least 60 days apart and free of artifact, and had height and weight measurements on both dates, were included in our longitudinal evaluations (n = 38 patients, n = 76 scans). In cases where a patient received numerous whole-body PET/CT scans on the same day, the first image collected that day or the CT corresponding to a concurrently imaged PET scan was used.

Healthy females, drawn from the *Healthy-Total-Body-CTs Dataset,*^26^ were adopted as a negative control sample for cachexia in cross-sectional comparisons. We included all female patients who had a skull-to-thigh CT scan free of artifact, in concert with body height and weight measurements obtained at the time of imaging (n = 16). The first full-body scan was used in any case where a patient received multiple scans on the same day.

To establish a positive control sample of cachexia for our cross-sectional and longitudinal comparisons, female patients recently diagnosed with stage IIB-III non-small cell lung cancer (NSCLC) were drawn from the ACRIN-NSCLC-FDG-PET dataset,^27^ which originated from the multi-center ACRIN 6668 clinical trial.^28^ Patients were imaged with PET/CT from skull-to-thigh prior to initiation of treatment (*pre-treatment*) and again following completion of chemoradiation (*post-treatment*; 12-16 weeks after radiation and ≥ 4 weeks after chemotherapy completion). We included female patients with an artifact-free scan and measures of body height and weight at the *pre-treatment* timepoint in our cross-sectional comparisons (n = 47). Further, patients imaged at both *pre-treatment* and *post-treatment* that were free of artifact, as well as both measures of height and weight, were included in longitudinal comparisons (n = 38 patients, n = 76 scans).

### Evaluation of Body Composition

The axial slice corresponding to the greatest visibility of left and right transverse processes of the L3 vertebrae was manually identified from each computed tomography (CT) scan and was subsequently utilized for body composition evaluation. Tissue segmentation was performed via Slice-O-Matic (version 6.6), according to the Alberta Protocol, in which region growing and manual tagging are used to segment skeletal muscle (SKM), intramuscular adipose (IMAT), visceral adipose (VAT) and subcutaneous adipose (SAT) tissues.^29^

In addition to the Alberta protocol, two further segmentation approaches of the skeletal muscle were performed. First, SKM and IMAT tissues from the Alberta protocol were combined into one summated tissue to represent all tissues within the muscle fascia and thus signify the entire muscular compartment, abbreviated as “M_COMP_”. Additionally, M_COMP_ volumes were further classified into six subclasses corresponding with tissue quality, each defined by a predetermined radiodensity range (in Hounsfield units, HU), similar to previously reported methods.^30–32^ Low density fat (LDF; -190 to -51 HU) and high density fat (HDF; -50 to -30 HU) combine to represent IMAT tissue, whereas very low density muscle (VLDM; -29 to -1 HU), low density muscle (LDM; 0 to 29 HU), normal density muscle (NDM; 30 to 100 HU), and high density muscle (101 to 150 HU) combine to represent SKM tissue. High density muscle (101 to 150 HU) has previously been suggested to reflect underlying bone tissue, and thus was not utilized.

Summary parameters, including cross-sectional surface area (CSA) and mean attenuation (HU), were determined for all segmented tissues from each approach, as well as combined to calculate relative proportional measures. In addition to merging SKM and IMAT into M_COMP_, VAT and SAT were also combined to represent major external adipose depots, abbreviated as “ExAT”. Similarly, all four soft tissues (SKM, IMAT, VAT, SAT) were combined to reflect total soft tissue quantified from the L3 axial slice, designated as “soft tissue”.

### Body Composition Calculations

Lean body mass (LBM) estimations were calculated using the James formula. Skeletal muscle index (SMI) was calculated as SKM (-29 to 150 HU) CSA of the L3 slice (cm^2^) divided by the patients squared body height (m^2^). Skeletal muscle gauge (SMG) was calculated as SMI multiplied by the average radiodensity of all muscle tissue (HU), as reported previously.^33^

Considering tissue quantity and attenuation are independent, though may be partially or conditionally dependent within or between participants (i.e. based on scan parameters, such as voxel size, etc), we sought to capture the multifaceted nature of tissues using a variable similar to SMG. As such, we also characterized tissues by the product of CSA (cm^2^) and average attenuation (HU) determined from the L3 slice, reported as CSA*HU; this combined evaluation of tissue quantity (e.g., CSA) and quality (e.g., attenuation, radiodensity) is intended to enhance the ability to capture longitudinal changes within a cohort by reflecting changes to a tissue’s quantity, quality, or both, which may vary between patients.

As is standard amid other methods of body composition evaluation, though less common in CT-derived estimations, proportional representations of tissues were also considered. Individual tissues’ CSA contributions towards total tissue within the muscle compartment (% of M_COMP_), regional deposition of external adipose tissues (% of ExAT), and total soft tissue (% of Soft Tissue), as measured from the L3 slice, were calculated as relative CSA proportions and expressed as “%Tissue” (i.e. %SKM, %NDM, etc).

Additional patient-specific calculations were performed to represent associations across a patients’ tissues, including CSA ratios and attenuation differences (HU diff). The CSA of various tissues were compared as ratios, including VAT/SAT, SKM/IMAT, and M_COMP_/ExAT ratios. Additionally, the average attenuation (in HU) of associated tissues were also compared as the calculated difference in attenuation between two tissues, including VAT-SAT and SKM-M_COMP_ HU differences.

### Characterizing Cancer Cachexia

Patients were characterized using defined criteria for cachexia using the traditional two-group model proposed by Fearon et al (2011).^4^ Differences in body mass between *pre*- and *post-treatment* were normalized to a six-month time frame for each patient. Individuals who exceeded a loss of 5% body weight, or a loss of 2% if they had a post-treatment BMI < 20 kg/m^2^, were classified as *cachectic*.

### Statistical Analyses

Statistical analyses were performed using JMP Pro (version 18.0). Data are reported as mean ± standard deviation (M±SD). While normality was considered, it was not used to determine statistical tests; rather, sample-specific residuals from mixed models were used to evaluate normality. Post-hoc evaluations of residuals were mostly considered to be normally distributed, as determined by the Shapiro-Wilk test.

All tests were performed as two-tailed, with significance determined as α<0.05. In intra-patient comparisons, terminology used to reference image or measurement timing relative to anticancer therapy is related to included samples as follows: *pre-NAC* and *post-NAC* terminology is used for the sole inclusion of patients with BC, while *pre-treatment* and *post-treatment* are used more generally to encompass various non-surgical treatments for both BC and NSCLC, though these two conventions are otherwise equivalent. Absolute and relative *pre-* to *post-treatment* changes were calculated for each patient (Δ = CT_Post-treatment_ - CT_Pre-treatment_) and were normalized to an inter-scan duration of six months (182.5 days) for standardization across the sample; as such, absolute (ABSΔ/6M) and relative (%Δ/6M) changes reported across a sample group include patient-wise adjustments for time, in which a negative change reflects a loss or decrease over time. Cohen’s *d* effect size was used to characterize *pre-* to *post-treatment* normalized change, in which ±0.2 was considered a small effect, ±0.5 was considered a medium effect, and ±0.8 was considered a large effect.

In consideration for the increased likelihood of type 1 error corresponding with the number of reported body composition outcome variables, the Benjamini-Hochberg procedure was used to apply a false discovery rate (FDR) correction with a threshold of 0.1 to one-way ANOVAs and fixed effects of mixed-effect models.^34^ Such p-values surviving FDR correction are reported with an asterisk (*).

#### Longitudinal Changes in Breast Cancer (Repeated Measures)

Mixed-effects models were performed using all *pre-* and *post-NAC* scans (n = 76 scans) from BC patients with scans at both timepoints (n = 38 patients). Body composition variables were evaluated across *time* (denoted as *pre-NAC* and *post-NAC* when exclusively referring to patients with BC), using inter-scan duration (i.e. days between scans) as a covariate and the patient identifiers as a random effect to limit influence from inherent differences across patients.

#### Multi-Sample Baseline Characterizations (Cross-sectional)

One-way ANOVAs were performed using measured and calculated body compositional variables across the three female patient *cohorts* (breast, healthy control, NSCLC cachexic control), including healthy (n = 16), BC at *pre-treatment* (n = 56), and NSCLC at *pre-treatment* (n = 47). No covariates were included in statistical models comparing pre-treatment body composition with the intention of encompassing any clinical hallmarks of naturally presenting disease states. Pairwise comparisons of specific groups were adjusted for multiple comparisons using Tukey’s HSD.

#### Longitudinal Monitoring of Females with Non-metastatic Cancer (Repeated Measures)

Mixed effect models were performed using all *pre*- and *post-treatment* scans (n = 152 scans) of BC and NSCLC patients imaged at both timepoints (n = 38 BC and n = 38 NSCLC, for a total of n = 76 patients) as a full factorial assessing the effects of *cohort* (BC, NSCLC) and the development of *cachexia* (non-cachectic, cachectic) across *time* (pre-treatment, post-treatment) with all interaction terms (IxD) included, using inter-scan duration as a covariate and patient identifiers as a random effect. Pairwise comparisons were adjusted for multiple comparisons using Tukey’s HSD and were performed for fixed effect IxD terms *Time*Cohort*, *Time*Cachexia*, and *Time*Cohort*Cachexia*. Additionally, the Wald Test was performed to characterize between subject differences, including the portion of model variance accounted for through inclusion of the patient as random effect.

## RESULTS

### Longitudinal Changes in Breast Cancer

To assess whether intra-patient changes to body composition occur in non-metastatic BC patients, we compared CT-derived body composition estimations from *pre-NAC* to *post-NAC* in patients recently diagnosed with stage II-III BC (n = 38), with respect to inter-scan duration. Demographics and clinical characteristics, as well as relative timing of patient scans, can be found in Table 1. Additionally, CT imaging parameters for this sample, as well as other samples utilized herein, can be found in Table S1.

**Table 1.** Breast Cancer Cohort Demographics & Clinical Characteristics.

| Variable |  | M ± SD or n | % |
| --- | --- | --- | --- |
| Total, *n* |  | 38 |  |
| Age at Dx |  | 51.9 ± 11.3 |  |
| Pre-Treatment Body Weight (kg) |  | 79.83 ± 13.81 |  |
| Pre-Treatment Body Height (cm) |  | 162.4 ± 6.2 |  |
| Pre-Treatment BMI (kg/m<sup>2</sup>) |  | 30.33 ± 5.48 |  |
| Pre-Treatment LBM (kg) |  | 48.58 ± 3.94 |  |
| Days from Dx to Pre-Treatment Scan |  | 35.8 ± 25.4 |  |
| Days from Dx to Surgery |  | 204.4 ± 43.8 (n = 36) |  |
| Days between scans |  | 147.6 ± 37.8 |  |
| ABS Weight Δ/6 months (kg) |  | -2.90 ± 4.63 |  |
| % Weight Δ/6 months (%) |  | -3.59 ± 5.51 |  |
| *Pre-Treatment BMI (n)* | Normal | 7 | 18.4% |
|  | Overweight | 13 | 34.2% |
|  | Obese I | 11 | 28.9% |
|  | Obese II | 5 | 13.2% |
|  | Obese III | 2 | 5.3% |
| *Tumor Subtype (n)* | BCL/Triple Negative | 11 | 28.9% |
|  | Luminal A | 15 | 39.5% |
|  | Luminal B | 5 | 13.2% |
|  | HER2 | 6 | 15.8% |
|  | Unknown | 1 | 2.6% |
| *ER Status (n)* | Positive | 21 | 55.3% |
|  | Negative | 17 | 44.7% |
| *PR Status (n)* | Positive | 15 | 39.5% |
|  | Negative | 23 | 60.5% |
| *HER2 Status (n)* | Positive | 11 | 28.9% |
|  | Negative | 26 | 68.4% |
|  | Unknown | 1 | 2.6% |
| *Menopausal Status (n)* | Pre-menopausal | 17 | 44.7% |
|  | Post-menopausal | 20 | 52.6% |
|  | Unknown | 1 | 2.6% |
| *Race (n)* | Asian | 1 | 2.6% |
|  | Black/African | 11 | 28.9% |
|  | White | 24 | 63.2% |
|  | Unknown | 2 | 5.3% |
| *Ethnicity (n)* | Hispanic or Latino | 1 | 2.6% |
|  | Non-Hispanic | 33 | 86.8% |
|  | Unknown | 4 | 10.5% |
| *Entry Disease stage (n)* | II | 15 | 39.5% |
|  | III | 23 | 60.5% |
| *ECOG Performance (n)* | Asymptomatic | 30 | 78.9% |
|  | Symptomatic, full ambulatory | 7 | 18.4% |
|  | Symptomatic, in bed <50% of day | 1 | 2.6% |
| *Procedure (n)* | Lumpectomy | 10 | 26.3% |
|  | Simple Mastectomy | 6 | 15.8% |
|  | Modified Radical Mastectomy | 17 | 44.7% |
|  | Other/Not performed | 5 | 13.2% |
| *Pathologic Response (n)* | Complete Response | 9 | 23.7% |
|  | Partial Response | 16 | 42.1% |
|  | No Response | 11 | 28.9% |
|  | Unknown | 2 | 5.3% |
| *Cachectic status (n)* | Non-cachectic | 24 | 63.2% |
|  | Cachectic | 14 | 36.8% |

Data presented as Mean ± Standard Deviation or sample size [*n*, (%)]. [BCL, basal cell like; BMI, body mass index; ECOG, Eastern Cooperative Oncology Group] \*may not add up to 100% due to rounding.

Table 2 includes *pre-NAC* tissue characteristics, as well as *pre*- to *post-NAC* changes indexed to six months’ time. BC patients demonstrated significant declines in body mass (-3.6 ± 5.5%, p = 0.022*), as well as total soft tissue CSA (-4.9 ± 15.9%, p = 0.039*) measured from the L3 slice.

**Table 2.** Initial and Dynamic Characterization of Tissue in BC Patients.

| Tissue | Pre-Treatment | ABS $\Delta/6M$ | % $\Delta/6M$ | *p* | *d* |
| --- | --- | --- | --- | --- | --- |
| Body weight (kg) | 79.83 ± 13.81 | -2.90 ± 4.63 | -3.59 ± 5.51 | **0.0007\*** | -0.89 |
| BMI (kg/m<sup>2</sup>) | 30.33 ± 5.48 | -1.11 ± 1.75 | -3.59 ± 5.51 | **0.0006\*** | -0.90 |
| LBM (kg) | 48.58 ± 3.94 | -0.53 ± 0.97 | -1.10 ± 1.97 | **0.0022\*** | -0.76 |
| SKM CSA (cm<sup>2</sup>) | 128.4 ± 23.06 | -4.63 ± 10.85 | -3.43 ± 9.20 | **0.0108\*** | -0.60 |
| IMAT CSA (cm<sup>2</sup>) | 22.40 ± 6.86 | 1.18 ± 3.41 | 5.64 ± 16.46 | 0.0760 | 0.49 |
| VAT CSA (cm<sup>2</sup>) | 119.0 ± 73.06 | -0.85 ± 24.91 | -0.91 ± 27.98 | 0.5787 | -0.05 |
| SAT CSA (cm<sup>2</sup>) | 271.4 ± 109.9 | -19.48 ± 60.02 | -7.17 ± 26.33 | **0.0340\*** | -0.46 |
| M<sub>COMP</sub> CSA (cm<sup>2</sup>) | 150.8 ± 22.39 | -3.43 ± 11.49 | -2.11 ± 8.25 | 0.0788 | -0.42 |
| ExAT CSA (cm<sup>2</sup>) | 390.4 ± 158.7 | -20.33 ± 69.55 | -6.65 ± 22.47 | 0.0500 | -0.41 |
| Soft Tissue CSA (cm<sup>2</sup>) | 541.2 ± 168.9 | -23.77 ± 75.27 | -4.87 ± 15.85 | **0.0388\*** | -0.45 |
| LDF CSA (cm<sup>2</sup>) | 13.17 ± 4.89 | 0.60 ± 2.30 | 4.72 ± 21.23 | 0.2626 | 0.37 |
| HDF CSA (cm<sup>2</sup>) | 9.23 ± 2.44 | 0.58 ± 1.55 | 7.71 ± 17.77 | **0.0222\*** | 0.53 |
| VLDM CSA (cm<sup>2</sup>) | 16.68 ± 5.23 | 0.39 ± 2.61 | 4.52 ± 16.25 | 0.4860 | 0.21 |
| LDM CSA (cm<sup>2</sup>) | 34.42 ± 9.37 | -0.19 ± 5.08 | 0.61 ± 13.09 | 0.9727 | -0.05 |
| NDM CSA (cm<sup>2</sup>) | 76.99 ± 24.36 | -4.70 ± 11.51 | -4.81 ± 17.75 | **0.0121\*** | -0.58 |
| NDM mean HU | 49.58 ± 3.26 | -0.72 ± 2.70 | | 0.0888 | -0.38 |
| SKM mean HU | 32.48 ± 7.01 | -1.39 ± 4.11 | | **0.0411\*** | -0.48 |
| M<sub>COMP</sub> mean HU | 18.79 ± 10.11 | -2.12 ± 5.10 | | **0.0151\*** | -0.59 |
| VAT mean HU | -95.37 ± 6.71 | 1.39 ± 5.88 | | 0.0835 | 0.33 |
| SAT mean HU | -105.2 ± 5.43 | 2.46 ± 4.72 | | **0.0009\*** | 0.74 |
| ExAT mean HU | -102.7 ± 5.12 | 2.34 ± 4.47 | | **0.0010\*** | 0.74 |
| NDM CSA*HU | 3863 ± 1367 | -300.1 ± 648.0 | -5.69 ± 20.83 | **0.0041\*** | -0.65 |
| SKM CSA*HU | 4239 ± 1397 | -321.8 ± 644.9 | -6.05 ± 18.37 | **0.0019\*** | -0.71 |
| M<sub>COMP</sub> CSA*HU | 2903 ± 1726 | -378.5 ± 721.5 | -4.70 ± 72.74 | **0.0019\*** | -0.74 |
| VAT CSA*HU | -11651 ± 7627 | 160.3 ± 2534 | -1.92 ± 29.99 | 0.4464 | 0.09 |
| SAT CSA*HU | -28800 ± 12960 | 2583 ± 6901 | -8.99 ± 27.21 | **0.0185\*** | 0.53 |
| SMI (cm<sup>2</sup>/m<sup>2</sup>) | 48.75 ± 8.78 | -1.79 ± 4.20 | -3.43 ± 9.20 | **0.0100\*** | -0.60 |
| SMG | 1612 ± 543.4 | -124.8 ± 252.6 | -6.05 ± 18.37 | **0.0019\*** | -0.70 |
| VAT/SAT CSA ratio | 0.447 ± 0.253 | 0.012 ± 0.151 | 9.65 ± 30.39 | 0.6195 | 0.11 |
| SKM/IMAT CSA ratio | 6.31 ± 2.37 | -0.47 ± 1.05 | -7.38 ± 16.32 | **0.0124\*** | -0.63 |
| M<sub>COMP</sub>/ExAT CSA ratio | 0.447 ± 0.187 | 0.055 ± 0.191 | 8.46 ± 25.59 | **0.0400\*** | 0.41 |
| SKM-M<sub>COMP</sub> HU difference | 13.68 ± 4.04 | 0.73 ± 1.91 | 5.66 ± 14.76 | **0.0440\*** | -0.54 |
| VAT-SAT HU difference | 9.82 ± 5.88 | 1.08 ± 5.23 | 2.92 ± 73.41 | 0.0711 | 0.29 |

Pre-NAC and six-month changes in cross-sectional surface area (CSA; cm<sup>2</sup>), mean attenuation (HU), tissue quantity-quality products (CSA\*HU; cm<sup>2</sup>\*HU) and additional body composition variables presented as Mean ± SD. P-values and Cohen's d effect sizes represent paired *pre-* to *post-NAC* differences among patients imaged at both timepoints (n=38), in which significant changes (*p*<0.05) are in bold and reported as absolute (ABS $\Delta/6M$ ) and relative (% $\Delta/6M$ ) change adjusted to six-months' time. Significant *pre-* to *post-NAC* changes that survived FDR correction ( $\leq 0.1$ ) are denoted by an asterisk (\*). [BMI, body mass index; ExAT, external adipose tissue; HDF, high density fat; HU, Hounsfield units; IMAT, intramuscular adipose tissue; LBM, lean body mass; LDF, low density fat; LDM, low density muscle; M<sub>COMP</sub>, muscle compartment tissues; NDM, normal density muscle; SAT, subcutaneous adipose tissue; SKM, skeletal muscle; SMG, skeletal muscle gauge; SMI, skeletal muscle index; VAT, visceral adipose tissue; VLDM, very low density muscle]

#### Tissue Characteristics

*Pre-* to *post-NAC* changes to tissues within the M_COMP_ suggest no significant change in tissue quantity, though a decrease in attenuation. When considering changes to tissue quantity and quality over time (i.e. CSA*HU), significant declines to quantity-quality products were observed across intra-compartmental tissues. (M_COMP_ CSA*HU p = 0.002*). Among the muscle compartments’ major constituents, there were no changes to IMAT CSA (p = 0.076), though significant decreases in both SKM CSA and attenuation were apparent, as well as in their combined product (SKM CSA*HU p = 0.002*). These intra-compartment changes to SKM and IMAT correspond with a significant decrease in SKM/IMAT CSA ratio in BC patients.

Further examination of tissues within the muscle compartment demonstrated a significant decrease in NDM CSA (-4.8 ± 17.8%, p = 0.012*), which constitutes a bulk of M_COMP_ tissue, as well as an increase in HDF CSA.

As for ExAT depots, there was a significant decrease in SAT CSA and an increase in SAT attenuation, however, no change in VAT CSA or attenuation.

#### Body and Muscle Composition

All *pre-NAC* tissue proportions, as well as *pre*- to *post-NAC* proportional changes, can be found in Table 3. Tissues’ relative contributions towards total soft tissue CSA were stable for %SKM and %VAT, as well as %ExAT and %M_COMP_; however, significant increases in soft tissue contributions from %IMAT and a relative decrease in contribution from %SAT were observed.

**Table 3.** Initial and Dynamic Tissue Proportions in BC Patients.

| Parent Tissue | Tissue | Pre-Treatment | ABS $\Delta$/6M | *p* | *d* |
| --- | --- | --- | --- | --- | --- |
| % of ExAT | VAT | 28.99 ± 11.19 | 1.08 ± 5.93 | 0.2202 | 0.26 |
| | SAT | 71.01 ± 11.19 | -1.08 ± 5.93 | 0.2202 | -0.26 |
| % of M<sub>COMP</sub> | SKM | 84.87 ± 4.84 | -1.15 ± 2.49 | **0.0057\*** | -0.66 |
| | IMAT | 15.13 ± 4.84 | 1.15 ± 2.49 | **0.0057\*** | 0.65 |
| | LDF | 8.93 ± 3.46 | 0.57 ± 1.58 | 0.0733 | 0.51 |
| | HDF | 6.20 ± 1.64 | 0.57 ± 1.19 | **0.0008\*** | 0.69 |
| | VLDM | 11.19 ± 3.45 | 0.58 ± 2.02 | 0.1148 | 0.41 |
| | LDM | 22.90 ± 5.41 | 0.43 ± 3.41 | 0.3785 | 0.18 |
| | NDM | 50.58 ± 12.13 | -2.09 ± 6.75 | 0.0677 | -0.44 |
| % of Soft Tissue | SKM | 25.59 ± 7.72 | 0.93 ± 4.56 | 0.1445 | 0.29 |
| | IMAT | 4.30 ± 1.12 | 0.58 ± 1.01 | **0.0005\*** | 0.81 |
| | M<sub>COMP</sub> | 29.89 ± 7.96 | 1.51 ± 5.21 | 0.0520* | 0.41 |
| | VAT | 20.70 ± 8.93 | 0.45 ± 4.13 | 0.5339 | 0.15 |
| | SAT | 49.41 ± 7.67 | -1.95 ± 6.15 | **0.0461\*** | -0.45 |
| | ExAT | 70.11 ± 7.97 | -1.51 ± 5.21 | 0.0520* | -0.41 |

*Pre-treatment* and six-month change (ABSΔ /6M) in relative tissue proportions (%) presented as Mean ± SD. P-values and Cohen's *d* effect sizes represent paired *pre-* to *post-NAC* differences among patients imaged at both timepoints (n=38), in which significant changes (*p*<0.05) are in bold and reported as absolute change in tissue contribution adjusted to six-months' time (ABSΔ /6M). *Pre-* to *post-NAC* changes that survived FDR correction (< 0.1) are denoted by an asterisk (\*). [ExAT, external adipose tissue; HDF, high density fat; IMAT, intramuscular adipose tissue; LDF, low density fat; LDM, low density muscle; M<sub>COMP</sub>, muscle compartment tissues; NDM, normal density muscle; SAT, subcutaneous adipose tissue; SKM, skeletal muscle; VAT, visceral adipose tissue; VLDM, very low density muscle]

As it pertains to tissues comprising the M_COMP_, proportional contributions from %SKM declined, alongside a proportional increase in contributions from %IMAT. Among precise M_COMP_ subclasses, a proportional increase was found only among %HDF, though proportional changes trended for most subclasses.

#### Descriptive Characterization of Tissue Changes

Figure 1 summarizes *pre-NAC* and six-month change profiles for each BC patient for body weight (Fig. 1A) and tissues traditionally segmented under the Alberta protocol (SKM, IMAT, VAT, SAT), expressed as CSA (Fig. 1B, 1D), as well as proportion of total soft tissue (Fig. 1C, 1E). There were no associations between six-month weight changes (Fig. 1A) and *pre-NAC* tissue profiles (Fig 1B-C). As demonstrated in Figure 1B, SKM CSA was fairly consistent across patients, with most variability in soft tissue CSA driven primarily by VAT and SAT tissues. Further, as seen in Fig. 1D, changes to total soft tissue corresponded with weight changes (ABS/6M: *r* = 0.631, R^2^ = 0.398, p < 0.0001; %/6M: *r* = 0.592, R^2^ = 0.350, p < 0.0001), and most tissues tended to change in a similar manner for a given patient. Importantly, the magnitude of CSA changes in VAT and SAT was much greater than that of SKM (Fig. 1D), indicating soft tissue changes are primarily driven by changes to ExAT tissues. These imbalanced changes across tissues leads to a substantially different interpretation when expressed as soft tissue proportions; as seen in Fig. 1E, these changes are reflected in many patients as an increase in SKM proportion, even for patients that actually had a decrease or no change to SKM CSA (ABS/6M %SKM of ST ∼ %/6M SKM CSA: *r* = 0.219, R^2^ = 0.048, p = 0.187).

**Figure 1A-E.**
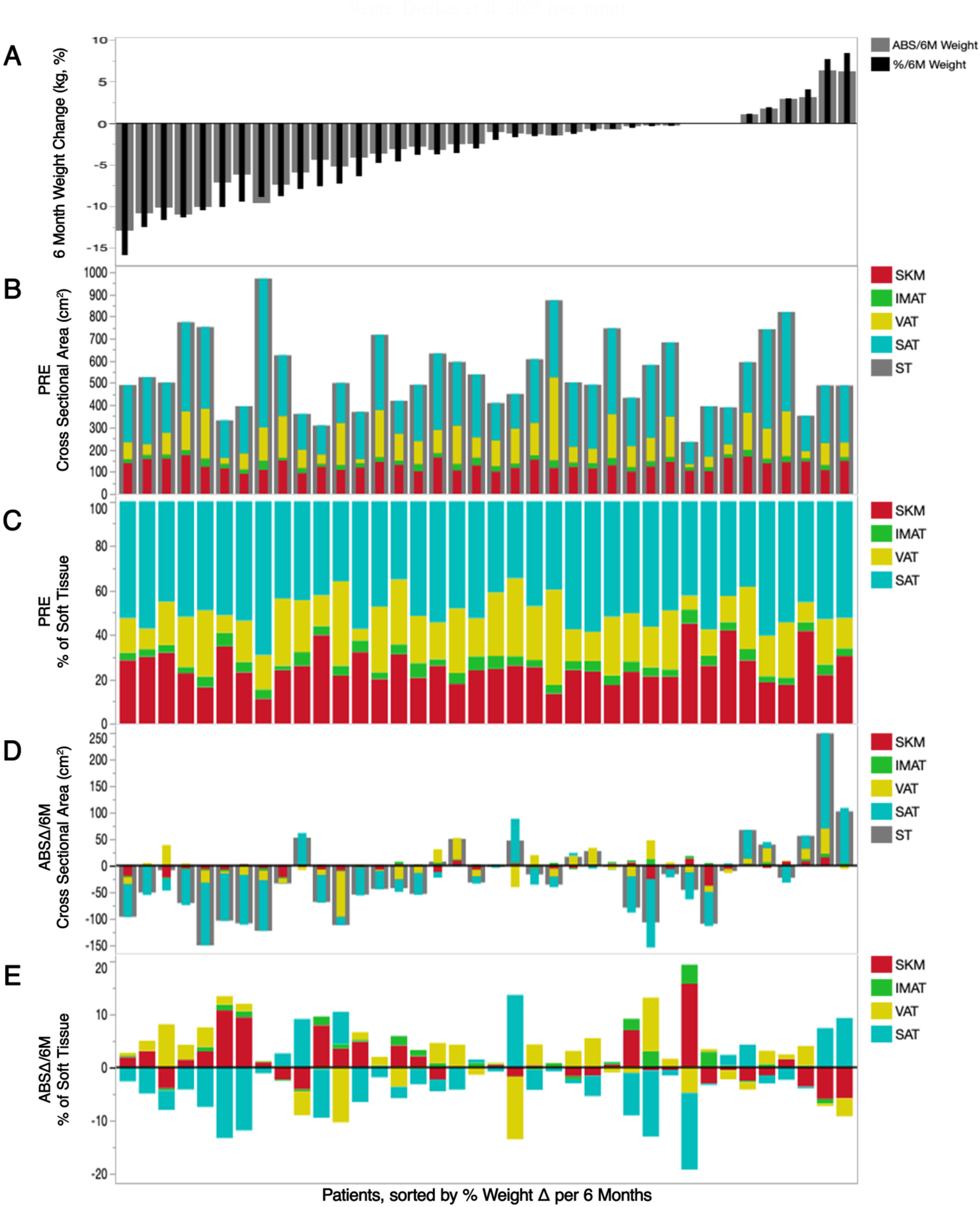
Patient Body Composition Profiles at Pre-Treatment and Throughout Treatment. Individual patient profiles at *pre-treatment* (B, C) and six-month changes (A, D, E) in body weight (A) and body composition (B-E) in patients with BC (n=38), in order of relative six-month changes (%Δ/6M) in body weight. A) absolute (ABSΔ/6M) and relative (%Δ/6M) changes in body weight (kg, %); B) cross-sectional area (CSA; in cm^2^) of tissues traditionally segmented under the Alberta protocol at *pre-treatment*; C) tissue percentage of total soft tissue CSA (%) at *pre-treatment*; D) ABSΔ/6M of tissue CSA (cm^2^); E) ABSΔ/6M of tissue proportions toward total soft tissue CSA (%). [IMAT, intramuscular adipose tissue; SAT, subcutaneous adipose tissue; SKM, skeletal muscle; ST, soft tissue; VAT, visceral adipose tissue]

Supplementary Figure S1 shows *pre-NAC* and six-month change profiles by patient for M_COMP_ tissues, including total CSA, subclass CSA, and subclass proportion of total M_COMP_. Nearly all patients demonstrated changes to intracompartmental tissues, and in most patients, substantially exceeded the magnitude of change measured to total M_COMP_ CSA (Fig. S1G). Importantly, 26 of the 38 patients with BC had a negative change in NDM CSA, which includes 7 patients that demonstrated an increase in total M_COMP_ CSA. Further, 13 patients had declines only in NDM concurrent with substantial increases among lower quality subclasses; another 11 patients demonstrated declines in both NDM and other intracompartmental subclasses, which was seen mostly in patients who lost substantial amounts of body weight. Therefore, a lack of change to total M_COMP_ CSA does not necessarily indicate a lack of change in the individual tissues that comprise the M_COMP_.

While significant declines were found only for M_COMP_ quality in the full sample, but not tissue quantity, divergent profiles regarding the manner of change(s) to muscle tissues existed across patients (Fig. 2). Approximately half the sample (n = 20, 52.6%) showed clinically significant declines in M_COMP_ quantity (Δ 2 cm^2^), and roughly half (n = 17, 44.7%) had clinically significant declines in M_COMP_ attenuation (≥ 2 HU), though these almost exclusively represent different patients. Only 5 of the 38 patients (13.2%) demonstrated declines that exceed both 2 cm^2^ in CSA and 2 HU in attenuation; however, an additional 15 patients (39.5%) had CSA declines alongside stable or improved M_COMP_ attenuation, while 12 patients (31.6%) had attenuation declines alongside stable or increased M_COMP_ CSA. Resultingly, only 6 of the 38 patients (15.8%) did not reflect a clinically significant decrement in M_COMP_ quantity (CSA) and/or quality (HU). The prevalence of these trends in muscle tissue can be seen in Figure 2. Importantly, changes to combined M_COMP_ CSA*HU did not correspond with clinically significant changes to individual tissue facets (i.e. SKM CSA, IMAT CSA, M_COMP_ CSA, M_COMP_ mean HU). Collectively, this figure further demonstrates that patients classified as cachectic according to body weight also mostly represent patients showing a proportional increase in lean body tissue (i.e. ABSΔ/6M %M_COMP_ of soft tissue). This ultimately leads to a false reflection of muscular improvements for a third of the sample, particularly patients with overt wasting.

**Figure 2.**
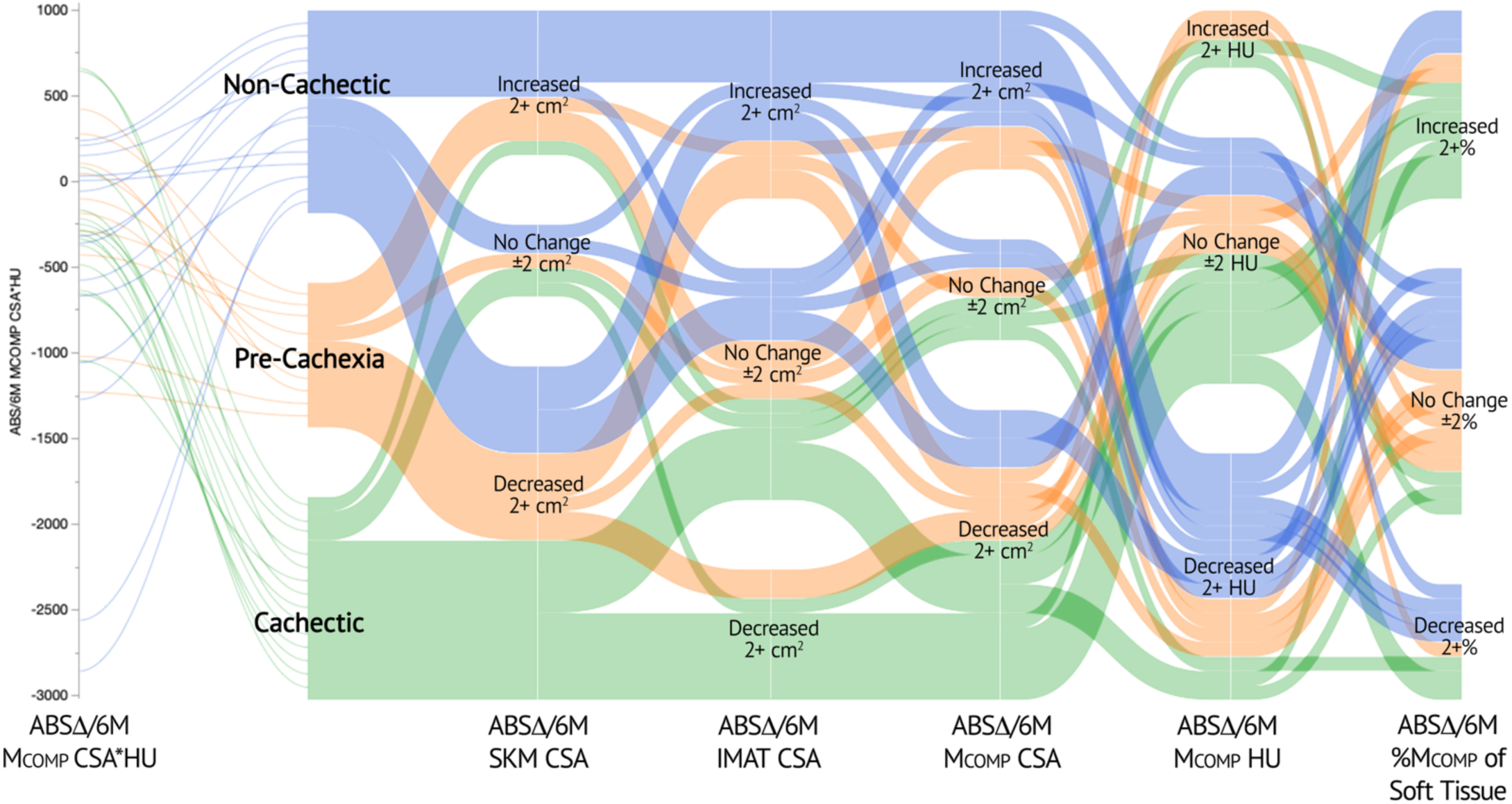
Individual Facets Contributing to Muscle Morphology in Patients with Breast Cancer. Summary of trends regarding muscle compartment (M_COMP_) changes in non-cachectic (n=14), pre-cachectic (n=10), and cachectic (n=14) patients with BC from *pre*- to *post-NAC*. [ABSΔ/6M, absolute change per six months; CSA, cross-sectional area; CSA*HU, product of tissue cross-sectional area and mean attenuation; HU, Hounsfield units; IMAT, intramuscular adipose tissue; SKM, skeletal muscle]

### Multi-Sample Baseline Characterizations

To evaluate baseline differences between patients with BC and other female cohorts with varying susceptibility to tissue wasting, *pre-treatment* body composition of BC patients (n = 56) was compared against healthy females (n = 16) and females with NSCLC prior to treatment initiation (n = 47). Clinical characteristics, anthropometrics, and segmented tissue properties for each cohort are provided in Table 4. The NSCLC cohort was significantly older than both the healthy (p < 0.001) and BC cohorts (p < 0.001). Further, despite no differences in body height, the BC patient cohort had a greater *pre-treatment* body mass (p < 0.001), BMI (p < 0.001), and LBM (p < 0.001) than the NSCLC patient cohort.

**Table 4.** Baseline Tissue Characteristics Across Cohorts.

| Variable | Healthy Controls | Breast Cancer | NSCLC | ANOVA, *p* |
| --- | --- | --- | --- | --- |
| Age <sup>b,c</sup> | 45.4 ± 11.2 | 51.7 ± 11.5 | 64.5 ± 8.7 | <0.0001* |
| Body mass (kg) <sup>c</sup> | 71.6 ± 14.7 | 80.4 ± 15.9 | 67.4 ± 14.4 | 0.0001* |
| Body height (cm) | 164.1 ± 4.8 | 163.1 ± 5.8 | 161.0 ± 5.7 | 0.0680* |
| BMI (kg/m<sup>2</sup>) <sup>c</sup> | 26.72 ± 6.14 | 30.23 ± 5.83 | 25.93 ± 5.00 | 0.0005* |
| LBM (kg) <sup>c</sup> | 47.07 ± 3.54 | 48.82 ± 4.16 | 45.28 ± 5.28 | 0.0007* |
| SKM CSA (cm<sup>2</sup>) <sup>b,c</sup> | 125.6 ± 12.37 | 128.5 ± 22.0 | 107.8 ± 17.96 | <0.0001* |
| IMAT CSA (cm<sup>2</sup>) <sup>b</sup> | 17.02 ± 7.01 | 23.06 ± 10.41 | 24.45 ± 9.65 | 0.0327* |
| VAT CSA (cm<sup>2</sup>) | 80.72 ± 61.30 | 114.4 ± 81.9 | 98.88 ± 66.07 | 0.2312 |
| SAT CSA (cm<sup>2</sup>) <sup>c</sup> | 224.3 ± 146.0 | 270.5 ± 121.4 | 194.6 ± 110 | 0.0073* |
| M<sub>COMP</sub> CSA (cm<sup>2</sup>) <sup>c</sup> | 142.6 ± 14.79 | 151.5 ± 24.69 | 132.2 ± 22.72 | 0.0002* |
| ExAT CSA (cm<sup>2</sup>) <sup>c</sup> | 305.1 ± 193.6 | 385.0 ± 173.2 | 293.5 ± 156.0 | 0.0195* |
| Soft Tissue CSA (cm<sup>2</sup>) <sup>c</sup> | 447.7 ± 203.7 | 536.5 ± 187.8 | 425.7 ± 171.1 | 0.0086* |
| LDF CSA (cm<sup>2</sup>) | 9.32 ± 4.87 | 13.37 ± 7.49 | 13.91 ± 6.57 | 0.0642* |
| HDF CSA (cm<sup>2</sup>) <sup>b</sup> | 7.69 ± 2.33 | 9.69 ± 3.35 | 10.54 ± 3.45 | 0.0129* |
| VLDM CSA (cm<sup>2</sup>) <sup>a,b</sup> | 12.83 ± 3.86 | 17.63 ± 7.25 | 18.32 ± 6.40 | 0.0153* |
| LDM CSA (cm<sup>2</sup>) <sup>a,b</sup> | 24.37 ± 8.38 | 34.89 ± 11.28 | 33.09 ± 9.59 | 0.0020* |
| NDM CSA (cm<sup>2</sup>) <sup>b,c</sup> | 88.12 ± 13.73 | 75.53 ± 23.20 | 55.72 ± 14.26 | <0.0001* |
| NDM mean HU <sup>a,b,c</sup> | 51.66 ± 3.11 | 49.56 ± 3.18 | 47.14 ± 2.58 | <0.0001* |
| SKM mean HU <sup>a,b,c</sup> | 38.54 ± 5.80 | 32.13 ± 7.58 | 27.85 ± 5.78 | <0.0001* |
| M<sub>COMP</sub> mean HU <sup>a,b,c</sup> | 27.50 ± 8.98 | 18.62 ± 11.04 | 12.42 ± 8.68 | <0.0001* |
| VAT mean HU | -89.54 ± 5.76 | -94.32 ± 6.82 | -91.37 ± 8.50 | 0.0334* |
| SAT mean HU <sup>c</sup> | -102.4 ± 2.66 | -105.2 ± 5.43 | -101.5 ± 8.23 | 0.0134* |
| ExAT mean HU <sup>c</sup> | -99.35 ± 2.36 | -102.5 ± 5.21 | -98.42 ± 7.91 | 0.0035* |
| NDM CSA*HU <sup>a,b,c</sup> | 4580 ± 900.4 | 3789 ± 1307 | 2646 ± 763.4 | <0.0001* |
| SKM CSA*HU <sup>b,c</sup> | 4839 ± 874.1 | 4163 ± 1328 | 3001 ± 805.1 | <0.0001* |
| M<sub>COMP</sub> CSA*HU <sup>a,b,c</sup> | 3870 ± 1196 | 2798 ± 1778 | 1580 ± 1181 | <0.0001* |
| VAT CSA*HU | -7518 ± 6054 | -11161 ± 8544 | -9413 ± 6598 | 0.1910 |
| SAT CSA*HU <sup>c</sup> | -22921 ± 14663 | -28771 ± 13960 | -20185 ± 12010 | 0.0054* |
| SMI (cm<sup>2</sup>/m<sup>2</sup>) <sup>b,c</sup> | 46.77 ± 5.68 | 48.33 ± 8.25 | 41.55 ± 6.38 | <0.0001* |
| SMG <sup>b,c</sup> | 1796 ± 315.4 | 1567 ± 506.2 | 1158 ± 310.7 | <0.0001* |
| VAT/SAT CSA ratio <sup>b</sup> | 0.355 ± 0.170 | 0.436 ± 0.267 | 0.544 ± 0.302 | 0.0295* |
| SKM/IMAT CSA ratio <sup>a,b,c</sup> | 8.61 ± 3.44 | 6.41 ± 2.49 | 4.92 ± 1.65 | <0.0001* |
| M<sub>COMP</sub>/ExAT CSA ratio | 0.661 ± 0.357 | 0.478 ± 0.252 | 0.629 ± 0.442 | 0.0497* |
| SKM- M<sub>COMP</sub> HU difference <sup>b</sup> | 11.04 ± 3.92 | 13.51 ± 4.65 | 15.43 ± 4.22 | 0.0021* |
| VAT-SAT HU difference | 12.81 ± 6.08 | 10.92 ± 5.83 | 10.14 ± 5.14 | 0.2607* |
| %SAT of ExAT <sup>b</sup> | 74.86 ± 8.82 | 71.73 ± 11.68 | 66.97 ± 11.92 | 0.0278* |
| %SKM of M<sub>COMP</sub> <sup>b,c</sup> | 88.23 ± 4.27 | 84.84 ± 5.49 | 81.77 ± 5.37 | 0.0001* |
| %LDF of M<sub>COMP</sub> <sup>b</sup> | 6.42 ± 3.04 | 8.77 ± 4.00 | 10.33 ± 3.93 | 0.0022* |
| %HDF of M<sub>COMP</sub> <sup>b,c</sup> | 5.35 ± 1.38 | 6.40 ± 1.77 | 7.91 ± 1.78 | <0.0001* |
| %VLDM of M<sub>COMP</sub> <sup>a,b,c</sup> | 8.92 ± 2.22 | 11.59 ± 3.94 | 13.72 ± 3.23 | <0.0001* |
| %LDM of M<sub>COMP</sub> <sup>a,b</sup> | 16.87 ± 4.82 | 22.91 ± 5.69 | 24.81 ± 4.29 | <0.0001* |
| %NDM of M<sub>COMP</sub> <sup>a,b,c</sup> | 62.23 ± 10.20 | 50.06 ± 13.33 | 42.75 ± 10.29 | <0.0001* |
| %SKM of Soft Tissue | 33.25 ± 12.74 | 26.40 ± 8.74 | 29.20 ± 11.54 | 0.0606* |
| %IMAT of Soft Tissue <sup>b,c</sup> | 3.96 ± 0.95 | 4.46 ± 1.49 | 6.19 ± 2.19 | <0.0001* |
| %M<sub>COMP</sub> of Soft Tissue | 37.21 ± 13.06 | 30.86 ± 9.16 | 35.39 ± 12.69 | 0.0488* |
| %VAT of Soft Tissue | 16.20 ± 7.67 | 19.85 ± 9.24 | 21.55 ± 9.27 | 0.1267 |
| %SAT of Soft Tissue <sup>c</sup> | 46.59 ± 9.59 | 49.29 ± 9.17 | 43.05 ± 10.67 | 0.0073* |

Cross-sectional surface area (cm<sup>2</sup>), mean attenuation (HU), tissue quantity-quality products (cm<sup>2</sup>\*HU), and other body composition variables presented as Mean ± SD. Significant groupwise differences (*p*<0.05) are denoted as follows: difference between a) healthy controls & breast cancer (BC), b) healthy controls & non-small cell lung cancer (NSCLC), c) BC & NSCLC. ANOVAs in which significance survived FDR correction (< 0.1) are denoted by an asterisk (\*). [BMI, body mass index; CSA, cross-sectional surface area; ExAT, external adipose tissue; HDF, high density fat; HU, Hounsfield units; IMAT, intramuscular adipose tissue; LBM, lean body mass; LDF, low density fat; LDM, low density muscle; M<sub>COMP</sub>, muscle compartment tissues; NDM, normal density muscle; SAT, subcutaneous adipose tissue; SKM, skeletal muscle; SMG, skeletal muscle gauge; SMI, skeletal muscle index; VAT, visceral adipose tissue; VLDM, very low density muscle]

#### Tissue Characteristics

*Pre-treatment* tissue quantities did not differ between healthy and BC patients for SKM, IMAT, M_COMP_, VAT, SAT, ExAT, or total soft tissue CSA. In contrast, BC patients had significantly greater CSA than did NSCLC patients across SKM (p < 0.001), M_COMP_ (p < 0.001), SAT (p = 0.005), ExAT (p = 0.020), and total soft tissue (p = 0.008).

Despite having comparable M_COMP_ tissue quantity (M_COMP_ CSA), BC patients had greater amounts of VLDM (p = 0.030) and LDM (p = 0.001) CSA than healthy patients, though they did not differ in LDF, HDF, or NDM CSA. In addition to having less overall tissue within the M_COMP_, NSCLC patients had less NDM tissue (NDM CSA p < 0.001) than BC patients, though were comparable in LDF, HDF, VLDM, and LDM CSA.

Cohort differences in the distribution of tissues that constitute the M_COMP_, independent of differences amid CSA, are further reflected by measures of M_COMP_ radiodensity and combined quantity-quality product; BC patients had significantly lower NDM (p = 0.035), SKM (p = 0.003) and M_COMP_ (p = 0.006) attenuations than healthy females, though still greater than females with NSCLC (NDM p < 0.001, SKM p = 0.005, M_COMP_ p = 0.006). Further, combined quantity-quality products for NDM and M_COMP_ also corresponded with expected disease-associated outcomes, in which BC patients had significantly lower CSA*HU than healthy (CSA*HU: NDM p = 0.028, M_COMP_ p = 0.034), though still greater than NSCLC patients (NDM and M_COMP_ CSA*HU p < 0.001).

These differences across cohorts to tissues within the M_COMP_ are illustrated in Figure 3. At higher HU values, BC patients had slightly less tissue than healthy females, though not significantly different in total area under the curve (i.e. NDM CSA). Comparatively, patients with BC were similar to NSCLC at lower HU values (i.e. LDF, HDF, VLDM, LDM).

**Figure 3A-B.**
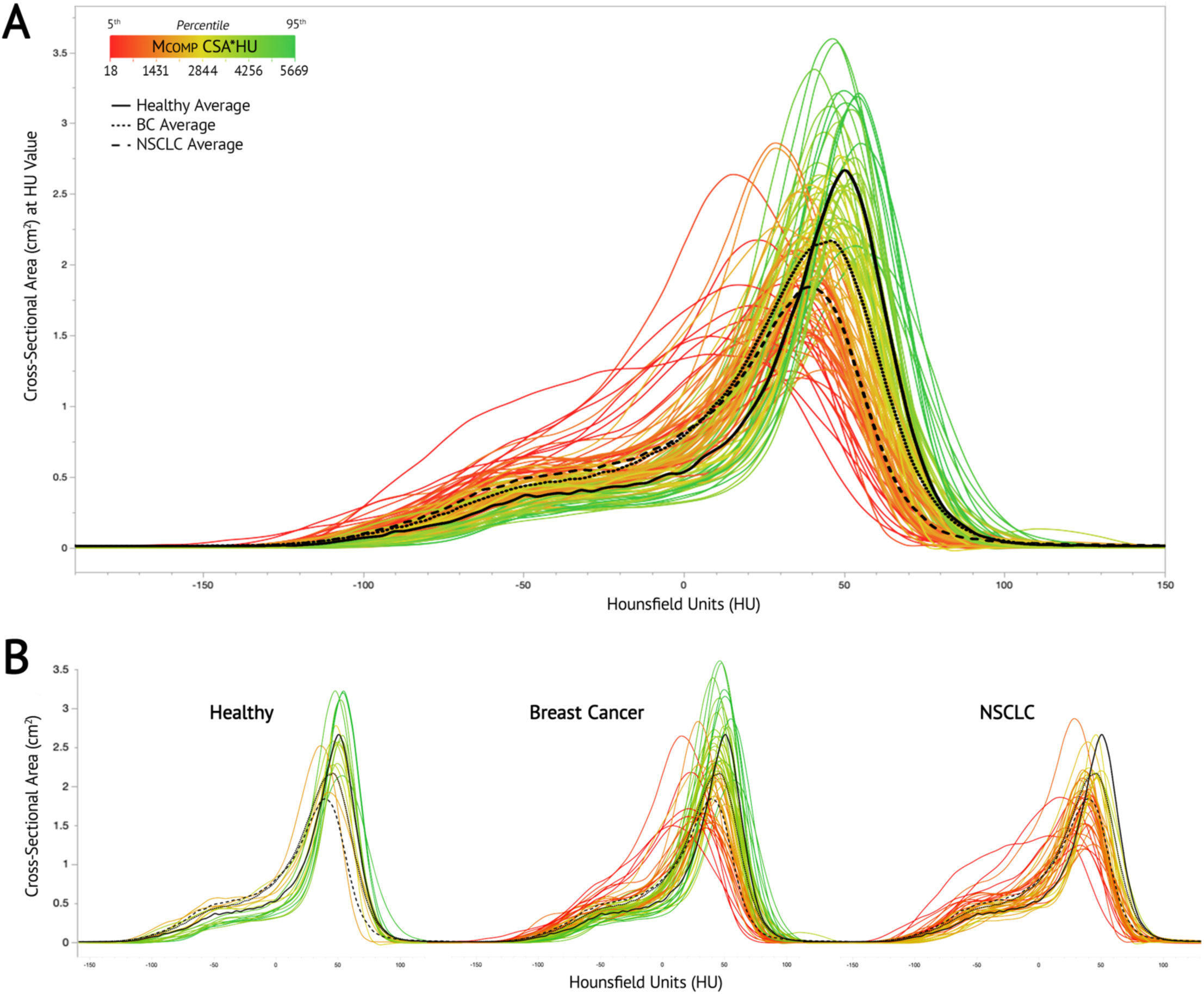
Characteristic Differences Across Cohorts in Muscle Compartment Tissues. Cross-sectional area of tissues within the muscle compartment (M_COMP_) by radiodensity for all healthy females (n=16) and female patients with BC (n=56) & NSCLC (n=47) at *pre-treatment*. Each curve represents a single patient, colored according to the quantity-quality product (CSA*HU) of their total M_COMP_ tissue. Comparison of M_COMP_ tissue distributions among A) all female samples (n=119), and B) by cohort.

No cohort differences existed for VAT attenuation, however, the BC cohort had significantly lower attenuations for SAT (p = 0.012) and ExAT depots (p = 0.003) than the NSCLC cohort.

#### Body and Muscle Composition

Proportional differences across cohorts are also reported in Table 4, including tissues’ relative contributions to ExAT depots, M_COMP_, and total soft tissue from the L3 slice. Minimal differences existed across cohorts in tissue contributions towards total soft tissue. NSCLC patients had a greater overall proportion of %IMAT tissue (6.2 ± 2.2%) than did both BC (4.5 ± 1.5%) and healthy (4.0 ± 1.0%) patients. Further, BC patients also had a significantly greater proportion of soft tissue derived from %SAT than NSCLC patients (p = 0.005). There were no differences between cohorts in soft tissue proportions from %SKM, %VAT, %M_COMP_, or %ExAT tissues, despite notable differences in tissue CSA.

Among ExAT, healthy patients had a greater relative amount deposited subcutaneously as compared to NSCLC patients (p = 0.049).

As for relative composition of M_COMP_, BC patients were similar to healthy patients in terms of total lean tissue content (%SKM of M_COMP_: Healthy 88.2 ± 4.3%, BC 84.8 ± 5.5%), while %SKM contributions were lower in NSCLC patients (81.8 ± 5.4%). More specifically, total M_COMP_ contributions from %NDM and %VLDM subclasses were significantly different across all three cohorts, with decreasing %NDM and increasing %VLDM proportions corresponding with expected worsening disease prognosis. Between these two distinctive subclasses exists %LDM, in which patients with BC and NSCLC have significantly greater proportions than healthy females. In contrast, contributions from %HDF were significantly greater in NSCLC than both healthy and BC patients, which represents tissue traditionally considered as IMAT just below the radiodensity lower limit of SKM.

### Longitudinal Monitoring of Females with Non-metastatic Cancer

To characterize whether tissue morphological patterns among patients diagnosed with BC differ from other malignancies in which cachexia is prevalent, we compared *pre-* and *post-treatment* body composition among females diagnosed with stage II-III BC (n = 38) and NSCLC (n = 38). Of these patients, 14 patients with BC (36.8%) and 21 with NSCLC (55.3%) went on to develop cachexia, as determined by trajectories in body weight recorded during NAC and chemoradiation treatment, respectively. Clinical characteristics for these patient groups are provided in Table 5. Six-month weight change did not differ among cachectic groups (BC -7.6 ± 3.3 kg, NSCLC -7.4 ± 3.6 kg, p = 0.999), however, more weight was gained by non-cachectic patients with NSCLC than BC (BC -0.2 ± 2.7 kg, NSCLC 2.5 ± 2.9 kg, p = 0.038). Further, *pre-treatment* body mass and LBM did not differ between non-cachectic and cachectic patients with BC; however, a lower body mass (p = 0.023) and LBM (p = 0.038) existed at *pre-treatment* in patients with NSCLC that would not develop cachexia, versus those that would.

**Table 5.** Clinical Characteristics of Cachectic and Non-Cachectic Patients by Cancer Type.

| Clinical Characteristic |  | BC<br>Non-Cachectic | BC<br>Cachectic | NSCLC<br>Non-Cachectic | NSCLC<br>Cachectic |
| --- | --- | --- | --- | --- | --- |
| n (% of total with like cancer type) |  | 24 (63.2%) | 14 (36.8%) | 17 (44.7%) | 21 (55.3%) |
| Age |  | 53.5 ± 9.2 <sup>c,d</sup> | 49.1 ± 14.2 <sup>c,d</sup> | 64.1 ± 7.4 <sup>a,b</sup> | 65.1 ± 9.6 <sup>a,b</sup> |
| Height (cm) |  | 161.9 ± 6.9 | 163.3 ± 4.7 | 159.2 ± 5.4 | 161.0 ± 5.5 |
| Pre-Treatment Body Mass (kg) |  | 80.52 ± 12.85 <sup>c</sup> | 78.66 ± 15.77 <sup>c</sup> | 58.89 ± 10.16 <sup>a,b,d</sup> | 71.68 ± 14.40 <sup>c</sup> |
| Pre-Treatment BMI (kg/m<sup>2</sup>) |  | 30.79 ± 5.14 <sup>c</sup> | 29.55 ± 6.14 <sup>c</sup> | 23.15 ± 3.36 <sup>a,b,d</sup> | 27.69 ± 5.60 <sup>c</sup> |
| Pre-Treatment LBM (kg) |  | 48.64 ± 4.09 <sup>c</sup> | 48.48 ± 3.83 <sup>c</sup> | 42.40 ± 4.86 <sup>a,b,d</sup> | 46.24 ± 4.35 <sup>c</sup> |
| Days Between Scans |  | 142.1 ± 38.6 | 157.0 ± 35.9 | 161.7 ± 18.5 | 161.1 ± 19.7 |
| ABSΔ Body Weight/6M (kg) |  | -0.17 ± 2.67 <sup>b,c,d</sup> | -7.59 ± 3.31 <sup>c,d</sup> | 2.54 ± 2.89 <sup>a,b,d</sup> | -7.42 ± 3.60 <sup>c,d</sup> |
| %Δ Body Weight/6M (%) |  | -0.26 ± 3.40 <sup>b,c,d</sup> | -9.30 ± 3.24 <sup>c,d</sup> | 4.34 ± 4.65 <sup>a,b,d</sup> | -10.17 ± 4.08 <sup>c,d</sup> |
| Disease Stage (n, %) | Stage 2 | 10 (26.3%) | 5 (13.2%) | 0 | 0 |
|  | Stage 3 | 14 (36.8%) | 9 (23.7%) | 17 (44.7%) | 21 (55.3%) |
| Age at Diagnosis (n, %) | 40s or younger | 8 (21.1%) | 8 (21.1%) | 0 | 2 (5.3%) |
|  | 50s | 9 (23.7%) | 3 (7.9%) | 3 (7.9%) | 4 (10.5%) |
|  | 60s | 6 (15.8%) | 2 (5.3%) | 11 (28.9%) | 7 (18.4%) |
|  | 70s or older | 1 (2.6%) | 1 (2.6%) | 3 (7.9%) | 8 (21.1%) |
| Pre-Treatment BMI Class (n, %) | Underweight | 0 | 0 | 1 (2.6%) | 1 (2.6%) |
|  | Normal | 3 (7.9%) | 4 (10.5%) | 12 (31.6%) | 5 (13.2%) |
|  | Overweight | 9 (23.7%) | 4 (10.5%) | 3 (7.9%) | 10 (26.3%) |
|  | Obese I | 8 (21.1%) | 3 (7.9%) | 1 (2.6%) | 4 (10.5%) |
|  | Obese II | 2 (5.3%) | 3 (7.9%) | 0 | 0 |
|  | Obese III | 2 (5.3%) | 0 | 0 | 1 (2.6%) |
| ABSΔ/6M Body Weight (n, %) | Gain 5+ lbs | 4 (10.5%) | 0 | 9 (23.7%) | 0 |
|  | +/- 5 lbs | 14 (36.8%) | 1 (2.6%) | 8 (21.1%) | 1 (2.6%) |
|  | Lost 5+ lbs | 6 (15.8%) | 13 (34.2%) | 0 | 20 (52.6%) |
| Net ABSΔ/6M Body Weight (n, %) | Gain 1+ kg | 6 (15.8%) | 0 | 12 (31.6%) | 0 |
|  | +/- 1 kg | 8 (21.1%) | 0 | 3 (7.9%) | 0 |
|  | Lost 1+ kg | 10 (26.3%) | 14 (36.8%) | 2 (5.3%) | 21 (55.3%) |
| Race (n, %) | Asian | 0 | 1 (2.6%) | 1 (2.6%) | 0 |
|  | Black/African | 10 (26.3%) | 1 (2.6%) | 0 | 1 (2.6%) |
|  | White | 13 (34.2%) | 11 (28.9%) | 16 (42.1%) | 20 (52.6%) |
|  | Unknown | 1 (2.6%) | 1 (2.6%) | 0 | 0 |
| Ethnicity (n, %) | Hispanic/Latino | 0 | 1 (2.6%) | 0 | 1 (2.6%) |
|  | Non-Hispanic | 21 (55.3%) | 12 (31.6%) | 17 (44.7%) | 18 (47.4%) |
|  | Unknown | 3 (7.9%) | 1 (2.6%) | 0 | 2 (5.3%) |

Data presented as Mean ± Standard Deviation, or sample size [n] and proportion (%) of all patients diagnosed with that respective cancer type (total n=38 patients each). Significant group-wise differences are denoted as follows: a) different from non-cachectic breast cancer (BC), b) different from cachectic BC, c) different from non-cachectic non-small cell lung cancer (NSCLC), and d) different from cachectic NSCLC. [ABSΔ/6M, absolute six-month change; BMI, body mass index; LBM, lean body mass]

While a seemingly larger proportion of NSCLC patients developed cachexia, although not significant, the criteria in which patients are classified is based upon relative changes to body mass. *Pre-treatment* body mass was 21.0% greater in patients with BC than NSCLC, though only 9.1% greater in LBM. As such, classifications for absolute changes in body mass are also provided in Table 5 according to various criteria, including those proposed by Paige et al. (2014),^35^ as well as by St Jeor et al (1997).^36^ Under both criteria, which utilize absolute thresholds of 1 kg or 5 lbs to characterize six-month weight loss, respectively, BC and NSCLC groups are comparable.

Figure 4 demonstrates associations between six-month changes in body mass with changes to tissue characteristics across all repeatedly imaged patients with BC and NSCLC (n = 76). There was no association with body mass for changes in NDM, and weight change was only weakly associated with changes in SKM quantity (Fig 4A). Combined changes in both the quantity and quality of M_COMP_ tissues also demonstrated no association with changes in body mass (r = -0.078, p = 0.502). In contrast, changes in body mass were strongly associated in a positive manner with quantity changes to ExAT depots (r = 0.646, R^2^ = 0.417, p < 0.0001) and total soft tissue (r = 0.673, R^2^ = 0.453, p < 0.0001), while negatively associated with combined change in quantity and attenuation among SAT (r= -0.624, R^2^ = 0.389, p < 0.0001) and ExAT (r= -0.671, R^2^ =0.450, p < 0.0001) (Fig. 4A).

**Figure 4A-C.**
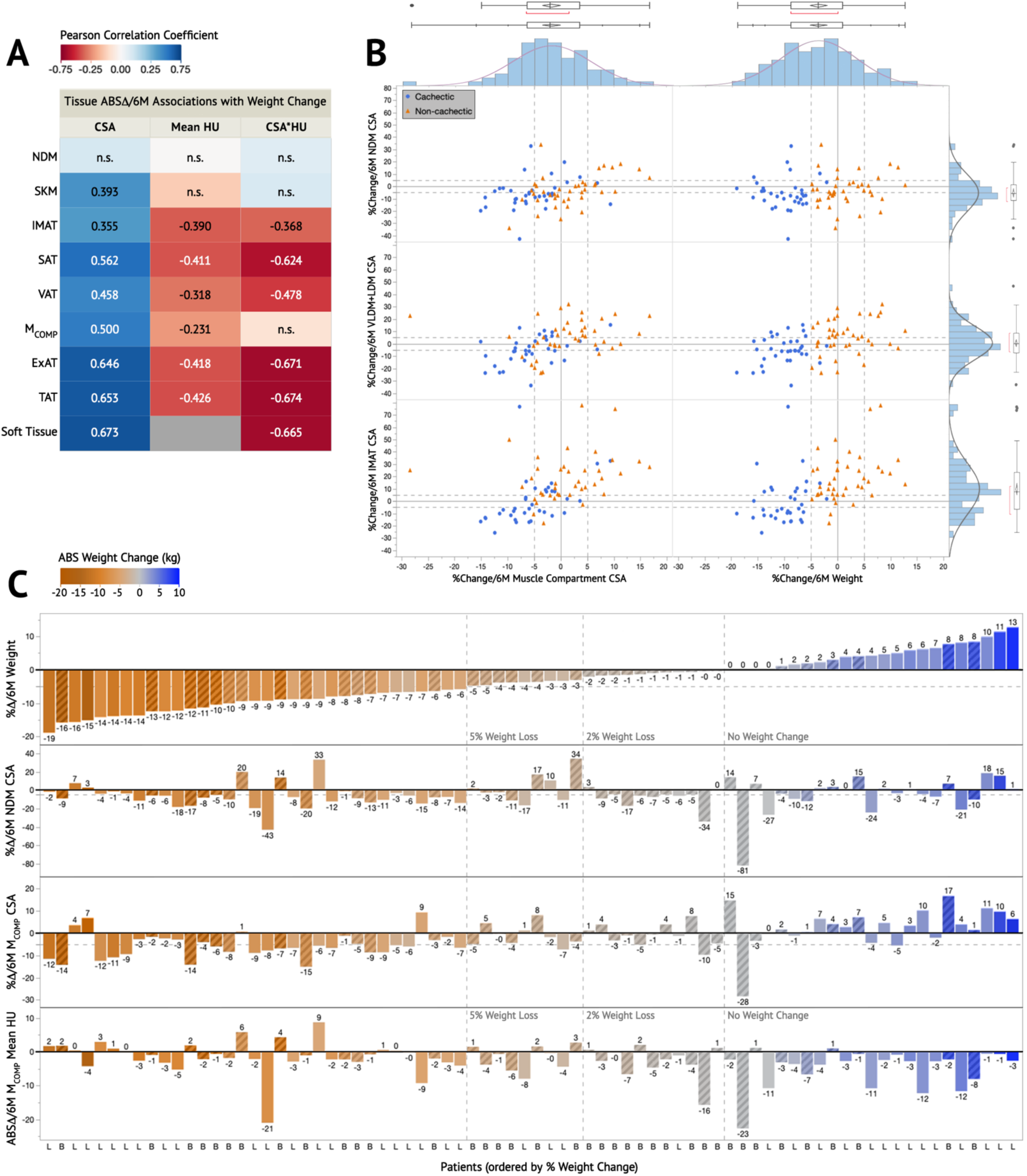
Longitudinal Associations Between Changes in Body Weight and Tissue Characteristics. Relationships between weight and body composition changes across six months in patients with BC (n=38) and NSCLC (n=38). A) Pearson’s correlations representing associations for absolute six-month changes (ABSΔ/6M) in body weight with ABSΔ/6M in the quantity (CSA), quality (mean attenuation) and quantity-quality products (CSA*HU) of various tissues. B) Comparison of normally distributed relative six-month changes (%Δ/6M) to muscle compartment (M_COMP_) CSA and body weight with *pre*- to *post-treatment* %Δ/6M in M_COMP_ tissue subclasses. C) Individual patient profiles of change as %Δ/6M body weight, %Δ/6M total M_COMP_ CSA, %Δ/6M NDM CSA, and ABSΔ/6M in M_COMP_ mean attenuation, ranked in order of %Δ/6M in body weight. Patients with BC are denoted with a “B” and striped bars, whereas NSCLC are denoted by an “L” and solid bars. [ABS, absolute; CSA, cross-sectional area; CSA*HU, product of tissue cross-sectional area and mean attenuation; ExAT, external adipose tissues; HU, Hounsfield units; IMAT, intramuscular adipose tissue; M_COMP_, muscle compartment; NDM, normal density muscle; SAT, subcutaneous adipose tissue; TAT, total adipose tissue; VAT, visceral adipose tissue; VLDM+LDM, sum of very low and low density muscle]

While the directionality of weight change tended to correspond with the directionality of change in M_COMP_ CSA (Fig. 4C), nearly all patients were characterized by declines in NDM CSA and M_COMP_ attenuation, independent of weight loss. As seen in Figure 4B, overt quantification of changes to total M_COMP_ CSA or body weight fail to delineate changes to muscle composition. Cachectic status had relatively no association with relative changes in NDM, with nearly all patients demonstrating a decline that exceeded 5% of their *pre-treatment* NDM CSA. Despite a lack of association with lean tissue, associations with cachectic status were strengthened with worsening tissue quality (Fig. 4B).

Linear mixed model statistics, including main effects of *time*, *cohort* (BC vs NSCLC), *cachectic development*, and all interaction terms, as well as statistics regarding between-patient variability, can be found in Table S2.

#### Cohort-Specific Changes

To evaluate whether longitudinal changes differ between patients with BC and NSCLC, the *Time*Cohort* interaction term was examined. Collectively, findings suggest NSCLC patients undergo more substantial changes throughout treatment pertaining to adipose within the M_COMP_ than do BC patients. More specifically, NSCLC patients were characterized by larger increases in IMAT CSA and %IMAT of M_COMP_, particularly LDF. This unique increase seen in NSCLC patients presented concurrent with declines in lean muscle (SKM CSA) across both cohorts, which resulted in significantly larger declines in M_COMP_ tissue quality (BC -2.1 ± 5.1 HU, NSCLC -3.3 ± 5.1 HU; p = 0.029), although this was lost following FDR correction. This was also reflected by a decline to the SKM/IMAT CSA ratio. Albeit, this change also manifested amid soft tissue proportions, with significantly larger increases in %IMAT contributions in NSCLC compared to BC.

Despite widespread *pre-treatment* differences across most tissues, as well as significant differences in age and BMI, longitudinal tissue changes were otherwise mostly similar between patients with BC and NSCLC. Similarities in changes to body tissues and muscle tissues across patients with BC and NSCLC can be seen in Supplementary Figure S2. Measured change among BC and NSCLC cohorts for all tissue parameters and relative tissue proportions can be found in Supplementary Table S3 and Table S4, respectively.

#### Changes with Cachexia Development

To evaluate whether longitudinal tissue dynamics differ between non-cachectic (n = 41) and cachectic (n = 35) patients, independent of cancer type, the *Time*Cachexia* interaction term was examined along with the corresponding pairwise comparisons. Collectively, findings suggest widespread differences in the manner and magnitude of *pre*- to *post-treatment* changes that correspond with the development of cachexia, despite no significant differences at either timepoint between non-cachectic and cachectic groups.

Table 6 includes *pre-treatment* and six-month changes to all tissue parameters for non-cachectic and cachectic patients, along with select fixed effect statistics regarding the *Time*Cachexia* interaction. Similar to changes in body weight, declines in soft tissue CSA occurred only in cachectic patients (non-cachectic 10.5 ± 72.4 cm^2^, p = 0.535; cachectic -70.4 ± 60.7 cm^2^, p < 0.001; *Time*Cachexia* p < 0.001*).

**Table 6.** Tissue Changes Throughout Non-Surgical Treatment in Cachectic and Non-Cachectic Patients.

| Tissue Parameter | Non-Cachectic |  |  |  | Cachectic |  |  |  | ME | ME | Time* |
| --- | --- | --- | --- | --- | --- | --- | --- | --- | --- | --- | --- |
|  | Pre-Treatment | ABSA/6M | *p* | *d* | Pre-Treatment | ABSA/6M | *p* | *d* | Time, *p* | Cachexia, *p* | Cachexia, *p* |
| Weight (kg) | 71.55 ± 15.89 | 0.96 ± 3.04 | 0.059 | 0.45 | 74.47 ± 15.14 | -7.49 ± 3.44 | <0.001 | -3.08 | <0.001* | 0.111 | <0.001* |
| BMI (kg/m<sup>2</sup>) | 27.62 ± 5.85 | 0.37 ± 1.17 | 0.062 | 0.44 | 28.43 ± 5.81 | -2.89 ± 1.34 | <0.001 | -3.04 | <0.001* | 0.220 | <0.001* |
| SKM CSA (cm<sup>2</sup>) | 118.9 ± 23.03 | -2.38 ± 9.67 | 0.380 | -0.35 | 114.2 ± 22.96 | -7.78 ± 7.13 | <0.001 | -1.54 | <0.001* | 0.947 | 0.002* |
| IMAT CSA (cm<sup>2</sup>) | 22.39 ± 6.59 | 3.56 ± 3.08 | <0.001 | 1.63 | 24.12 ± 10.12 | 0.31 ± 4.75 | 0.999 | 0.09 | <0.001* | 0.651 | <0.001* |
| VAT CSA (cm<sup>2</sup>) | 109.6 ± 73.45 | 10.94 ± 22.70 | 0.044 | 0.68 | 104.5 ± 62.88 | -16.58 ± 33.21 | 0.006 | -0.71 | 0.497 | 0.807 | <0.001* |
| SAT CSA (cm<sup>2</sup>) | 221.8 ± 104.9 | -1.63 ± 57.39 | 1.000 | -0.04 | 237.8 ± 128.5 | -46.39 ± 45.04 | <0.001 | -1.46 | <0.001* | 0.271 | <0.001* |
| M<sub>COMP</sub> CSA (cm<sup>2</sup>) | 141.3 ± 23.64 | 1.19 ± 9.74 | 0.575 | 0.17 | 138.3 ± 24.08 | -7.45 ± 8.30 | <0.001 | -1.27 | 0.003* | 0.910 | <0.001* |
| ExAT CSA (cm<sup>2</sup>) | 331.4 ± 160.6 | 9.30 ± 67.15 | 0.613 | 0.20 | 342.3 ± 167.7 | -62.97 ± 61.45 | <0.001 | -1.45 | 0.001* | 0.511 | <0.001* |
| Soft tissue CSA (cm<sup>2</sup>) | 472.7 ± 174.4 | 10.48 ± 72.37 | 0.535 | 0.21 | 480.7 ± 182.0 | -70.44 ± 60.73 | <0.001 | -1.64 | <0.001* | 0.532 | <0.001* |
| LDF CSA (cm<sup>2</sup>) | 12.94 ± 4.95 | 2.65 ± 2.14 | <0.001 | 1.75 | 14.09 ± 6.81 | 0.23 ± 3.66 | 1.000 | 0.09 | <0.001* | 0.636 | <0.001* |
| HDF CSA (cm<sup>2</sup>) | 9.46 ± 2.16 | 0.91 ± 1.51 | 0.002 | 0.85 | 10.03 ± 3.60 | 0.08 ± 1.55 | 0.996 | 0.08 | 0.009* | 0.733 | 0.023* |
| VLDM CSA (cm<sup>2</sup>) | 16.44 ± 4.36 | 1.06 ± 2.32 | 0.051 | 0.65 | 17.86 ± 6.15 | -0.48 ± 2.76 | 0.587 | -0.25 | 0.404 | 0.383 | 0.008* |
| LDM CSA (cm<sup>2</sup>) | 32.91 ± 9.67 | 0.85 ± 5.23 | 0.381 | 0.23 | 33.33 ± 7.23 | -2.32 ± 3.54 | 0.023 | -0.93 | 0.288 | 0.824 | 0.002* |
| NDM CSA (cm<sup>2</sup>) | 69.09 ± 21.89 | -4.03 ± 11.28 | 0.058 | -0.51 | 62.55 ± 24.01 | -4.77 ± 8.58 | 0.027 | -0.79 | <0.001* | 0.703 | 0.705 |
| NDM mean HU | 48.43 ± 3.31 | -0.44 ± 2.94 | 0.856 | -0.21 | 48.35 ± 3.12 | -0.33 ± 1.99 | 0.794 | -0.24 | 0.228 | 0.541 | 0.889 |
| SKM mean HU | 31.22 ± 6.30 | -1.81 ± 4.29 | 0.021 | -0.60 | 29.23 ± 7.21 | -0.62 ± 3.23 | 0.781 | -0.27 | 0.009* | 0.537 | 0.193 |
| M<sub>COMP</sub> mean HU | 16.99 ± 8.96 | -3.95 ± 5.17 | <0.001 | -1.08 | 14.09 ± 10.97 | -1.29 ± 4.70 | 0.493 | -0.39 | <0.001* | 0.583 | 0.015* |
| VAT mean HU | -93.89 ± 8.24 | -0.91 ± 6.52 | 0.269 | -0.20 | -93.05 ± 7.86 | 2.69 ± 4.94 | 0.011 | 0.77 | 0.272 | 0.764 | 0.001* |
| SAT mean HU | -103.4 ± 6.38 | 0.13 ± 4.72 | 0.998 | 0.04 | -103.8 ± 8.61 | 4.56 ± 5.96 | <0.001 | 1.08 | <0.001* | 0.689 | <0.001* |
| ExAT mean HU | -100.7 ± 6.49 | 0.04 ± 4.42 | 0.986 | 0.01 | -100.7 ± 8.12 | 4.23 ± 5.71 | <0.001 | 1.05 | <0.001* | 0.745 | <0.001* |
| NDM CSA*HU | 3396 ± 1241 | -241.5 ± 663.7 | 0.065 | -0.51 | 3068 ± 1321 | -261.5 ± 475.1 | 0.034 | -0.78 | <0.001* | 0.819 | 0.731 |
| SKM CSA*HU | 3770 ± 1263 | -279.2 ± 654.0 | 0.017 | -0.60 | 3417 ± 1368 | -318.0 ± 465.9 | 0.006 | -0.97 | <0.001* | 0.756 | 0.645 |
| M<sub>COMP</sub> CSA*HU | 2457 ± 1483 | -510.8 ± 723.5 | <0.001 | -1.0 | 1984 ± 1786 | -330.9 ± 675.5 | 0.050 | -0.69 | <0.001* | 0.647 | 0.245 |
| VAT CSA*HU | -10712 ± 7754 | -1125 ± 2440 | 0.042 | -0.65 | -10017 ± 6224 | 1717 ± 3336 | 0.005 | 0.73 | 0.469 | 0.737 | <0.001* |
| SAT CSA*HU | -23234 ± 11365 | 197.5 ± 5995 | 1.000 | 0.05 | -25298 ± 15234 | 5696 ± 5652 | <0.001 | 1.43 | <0.001* | 0.242 | <0.001* |
| SMI (cm<sup>2</sup>/m<sup>2</sup>) | 45.91 ± 8.11 | -0.96 ± 3.79 | 0.347 | -0.36 | 43.69 ± 9.15 | -2.98 ± 2.77 | <0.001 | -1.52 | <0.001* | 0.662 | 0.003* |
| SMG | 1454 ± 474.4 | -110.0 ± 257.9 | 0.017 | -0.60 | 1305 ± 528.2 | -121.5 ± 182.0 | 0.007 | -0.94 | <0.001* | 0.616 | 0.693 |
| VAT/SAT CSA ratio | 0.527 ± 0.311 | 0.064 ± 0.265 | 0.140 | 0.34 | 0.467 ± 0.244 | 0.009 ± 0.158 | 0.978 | 0.08 | 0.082* | 0.174 | 0.242 |
| SKM/IMAT CSA ratio | 5.77 ± 2.03 | -1.029 ± 1.223 | <0.001 | -1.19 | 5.43 ± 2.30 | -0.141 ± 0.871 | 0.864 | -0.23 | <0.001* | 0.902 | <0.001* |
| M<sub>COMP</sub>/ExAT CSA ratio | 0.555 ± 0.332 | -0.014 ± 0.244 | 0.760 | -0.08 | 0.543 ± 0.423 | 0.165 ± 0.301 | 0.003 | 0.78 | 0.050* | 0.749 | 0.001* |
| SKM-M<sub>COMP</sub> HU diff | 14.23 ± 4.01 | 2.14 ± 2.05 | <0.001 | 1.48 | 15.13 ± 4.66 | 0.67 ± 2.51 | 0.413 | 0.38 | <0.001* | 0.768 | 0.001* |
| VAT-SAT HU diff | 9.52 ± 5.44 | 1.04 ± 5.26 | 0.216 | 0.28 | 10.71 ± 5.27 | 1.87 ± 3.86 | 0.036 | 0.68 | 0.001* | 0.304 | 0.476 |

Pre-treatment and absolute six-month change (ABSA/6M) in cross-sectional surface area (cm<sup>2</sup>), mean attenuation (HU), tissue quantity-quality products (cm<sup>2</sup>\*HU), and other tissue parameters presented as Mean ± SD. No significant *Timepoint*\**Cachexia* differences (*p*<0.05) existed at *pre-treatment* or *post-treatment* between non-cachectic & cachectic patients. Fixed effects in which significance survived FDR correction (< 0.1) are denoted by an asterisk (\*). [BMI, body mass index; CSA, cross-sectional surface area; diff, difference; ExAT, external adipose tissue; HDF, high density fat; HU, Hounsfield units; IMAT, intramuscular adipose tissue; LBM, lean body mass; LDF, low density fat; LDM, low density muscle; M<sub>COMP</sub>, muscle compartment tissues; NDM, normal density muscle; SAT, subcutaneous adipose tissue; SKM, skeletal muscle; SMG, skeletal muscle gauge; SMI, skeletal muscle index; VAT, visceral adipose tissue; VLDM, very low density muscle]

Among tissue quantities, NDM CSA was the only tissue that did not exhibit a significant *Time*Cachexia* interaction (p = 0.705), in which average declines were similar for non-cachectic (-4.0 ± 11.3 cm^2^, p = 0.058) and cachectic (-4.8 ± 8.6 cm^2^, p = 0.027) patients. NDM attenuation, in which neither group experienced changes, and NDM CSA*HU, in which changes for both groups approached or reached significance, were also among the few other variables in which change over time did not differ between non-cachectic and cachectic patients.

Significant declines in SKM CSA were unique to cachectic patients. Conversely, declines to SKM attenuation and increases in IMAT CSA were unique to non-cachectic patients. Put together, significant declines to M_COMP_ quantity are unique to cachectic patients (non-cachectic 1.2 ± 9.7 cm^2^, p = 0.575; cachectic -7.5 ± 8.3 cm^2^, p < 0.001; *Time*Cachexia* p < 0.001*), whereas declines to M_COMP_ quality are unique to non-cachectic patients (non-cachectic -4.0 ± 5.2 HU, p < 0.001; cachectic -1.3 ± 4.7 HU, p = 0.493; *Time*Cachexia* p = 0.015*). Combined change to both quantity and quality, as reflected by M_COMP_ CSA*HU, suggests average declines among both patient groups of a comparable degree.

Significant decreases in CSA and increases in attenuation of adipose tissues, including VAT, SAT and ExAT, occurred only in cachectic patients. *Pre-* to *post-treatment* changes in CSA and attenuation of various tissues are shown in Figure 5.

**Figure 5A-E.**
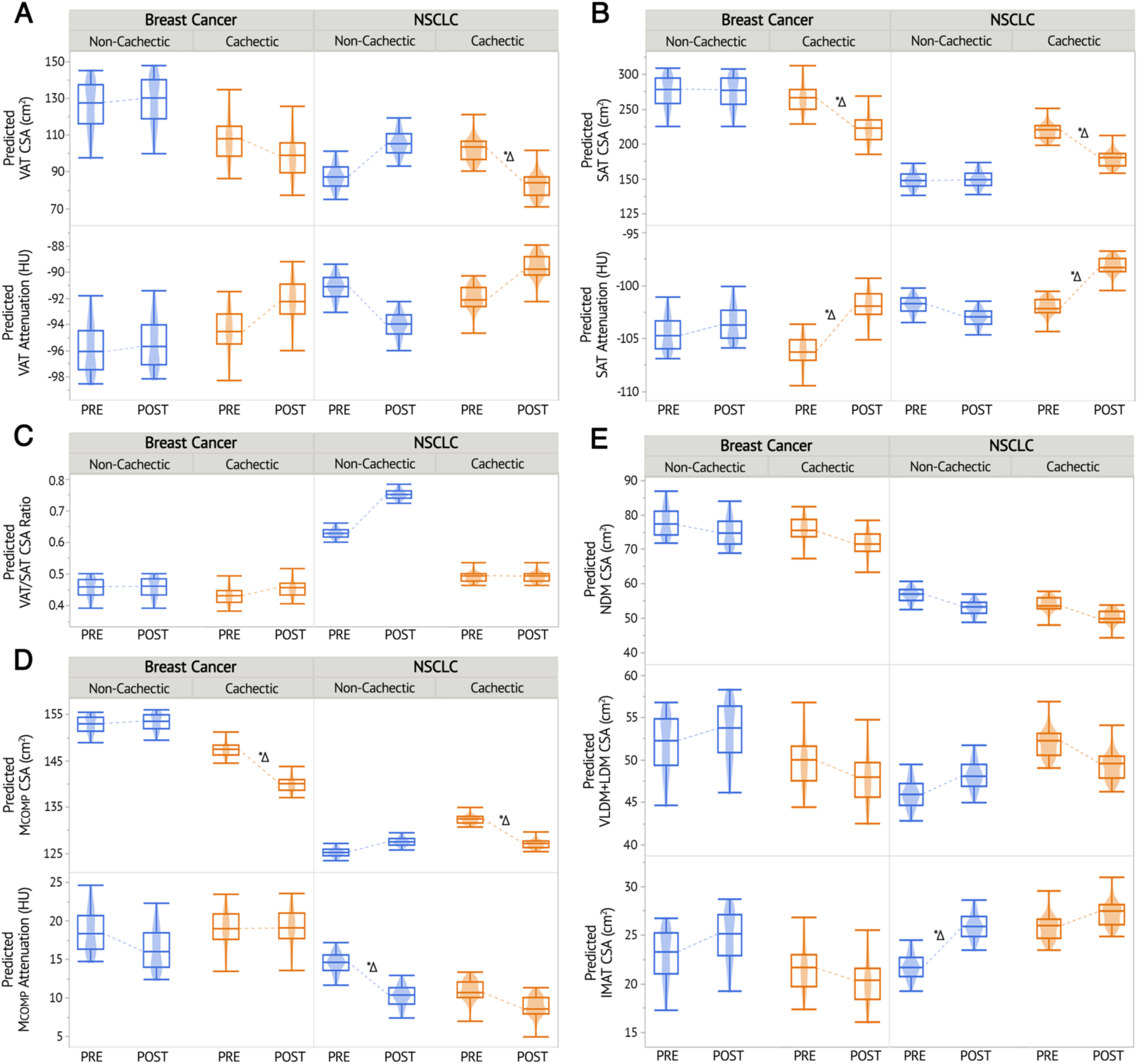
Muscle and Adipose Tissue Changes with Cachexia Development. Mixed model predicted values for CSA (cm^2^) & mean attenuation (HU) of A) visceral adipose (VAT), and B) subcutaneous adipose (SAT) tissues, C) predicted values for VAT/SAT CSA ratio, D) predicted values for CSA (cm^2^) & mean attenuation (HU) of muscle compartment (M_COMP_) tissues, E) predicted values for CSA area (cm^2^) of M_COMP_ tissue subclasses. Significant *pre*- to *post-treatment* pairwise comparisons (*p*<0.05) are denoted by *\*Δ*. [CSA, cross-sectional area; HU, Hounsfield units; IMAT, intramuscular adipose tissue; NDM, normal density muscle; VLDM+LDM, sum of very low and low density muscle]

Shifts to tissue proportions both within M_COMP_ and to ExAT depots occurred only in the non-cachectic group, in which there was a decrease in lean %SKM content (%SKM of M_COMP_: non-cachectic -2.5 ± 2.5%, p < 0.001; cachectic -1.0 ± 2.8%, p = 0.133; *Time*Cachexia* p = 0.005*), and a decrease in subcutaneously deposited ExAT (%SAT of ExAT: non-cachectic -2.6 ± 7.8%, p = 0.038; cachectic -0.7 ± 7.2%, p = 0.841; *Time*Cachexia* p = 0.219*). Supplementary Table S5 includes *pre-treatment* and six-month changes to all tissue proportions for non-cachectic and cachectic patients, along with select fixed effect statistics regarding the *Time*Cachexia* interaction. *Pre-* to *post-treatment* tissue proportions representing body and muscle composition are seen in Figure 6.

**Figure 6A-B.**
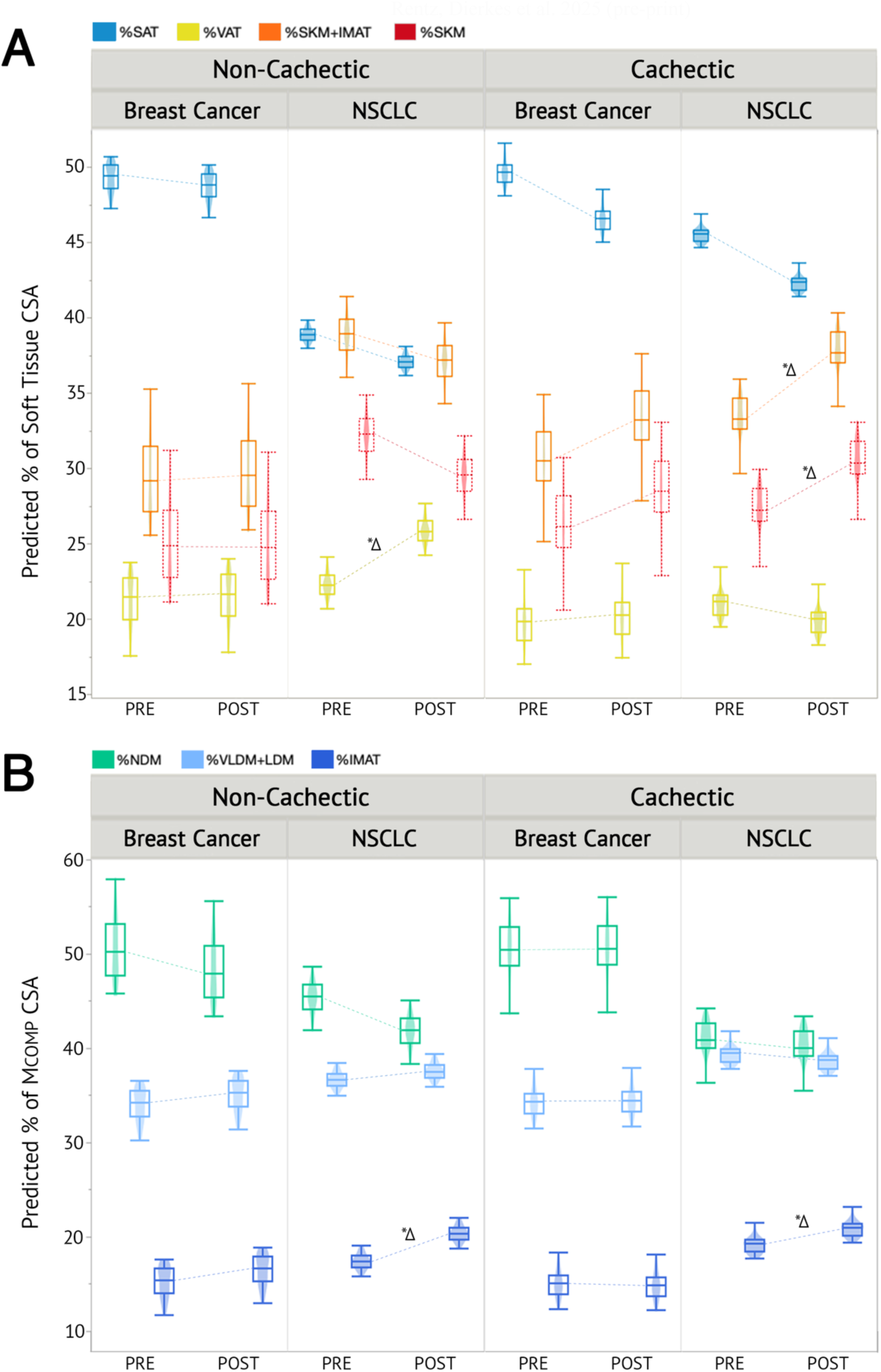
Proportional Changes in Body and Muscle Composition with Cachexia Development. Mixed model predicted values for tissue proportions of A) total soft tissue CSA (%), and B) total muscle compartment (M_COMP_) CSA. Significant *pre*- to *post-treatment* pairwise comparisons (*p*<0.05) are denoted by *\*Δ*. [CSA, cross-sectional area; IMAT, intramuscular adipose tissue; M_COMP_, muscle compartment; NDM, normal density muscle; SAT, subcutaneous adipose tissue; SKM, skeletal muscle; VAT, visceral adipose tissue; VLDM+LDM, sum of very low and low density muscle]

#### Differential Trajectories in Cachexia Development Across Cancer Types

To determine whether female patients with BC and NSCLC differ in tissue morphology during the development of cachexia, *pre*- to *post-treatment* changes were compared across cancer types and cachectic status (*Time*Cohort*Cachexia* interaction term). Dynamic changes captured throughout treatment to tissue characteristics and proportions are provided in Tables 7 and 8, respectively, based on the development of cachexia in patients with BC and NSCLC.

**Table 7.**
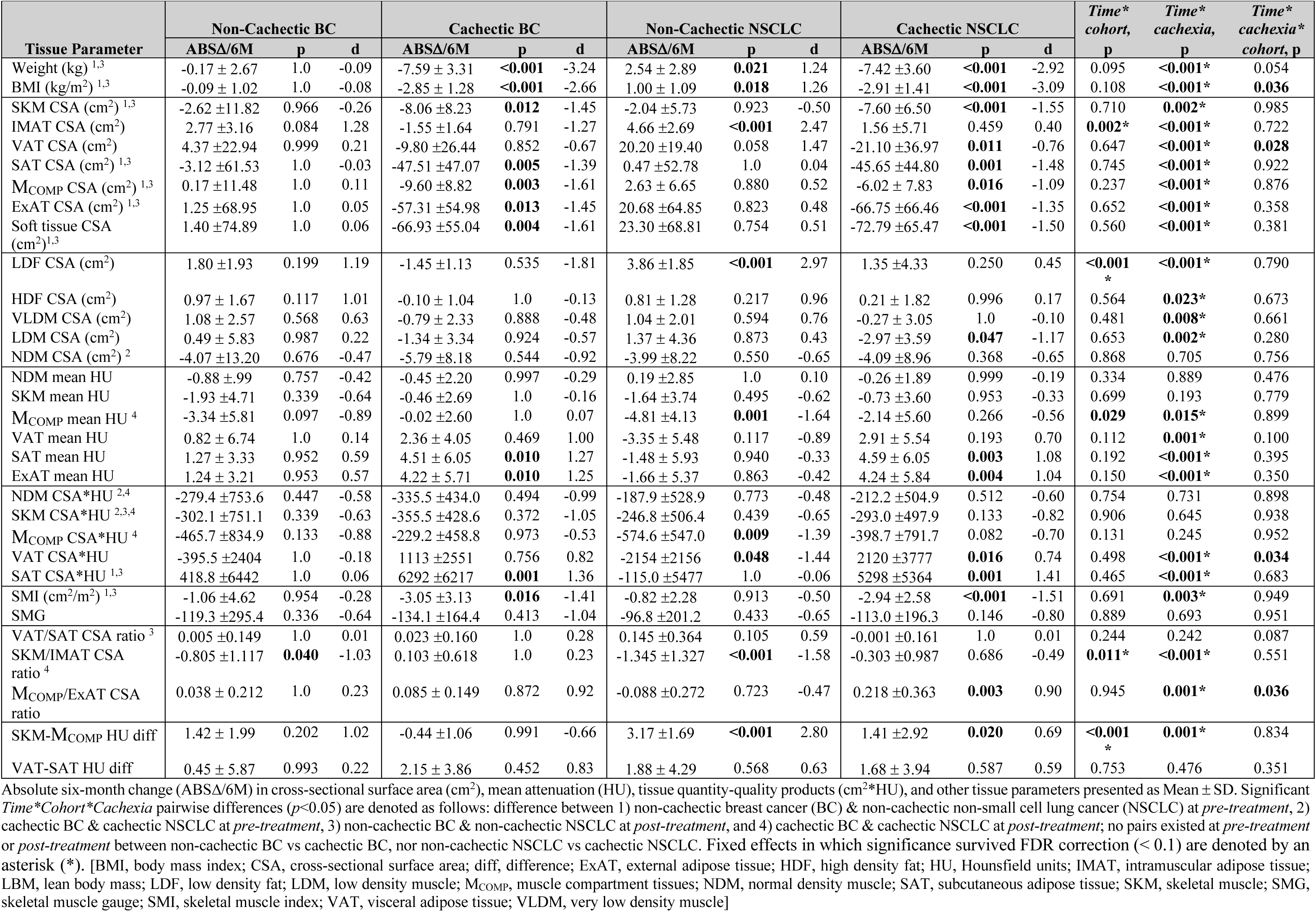
Tissue Dynamics During Cachexia Development by Cancer Type.

**Table 8.**
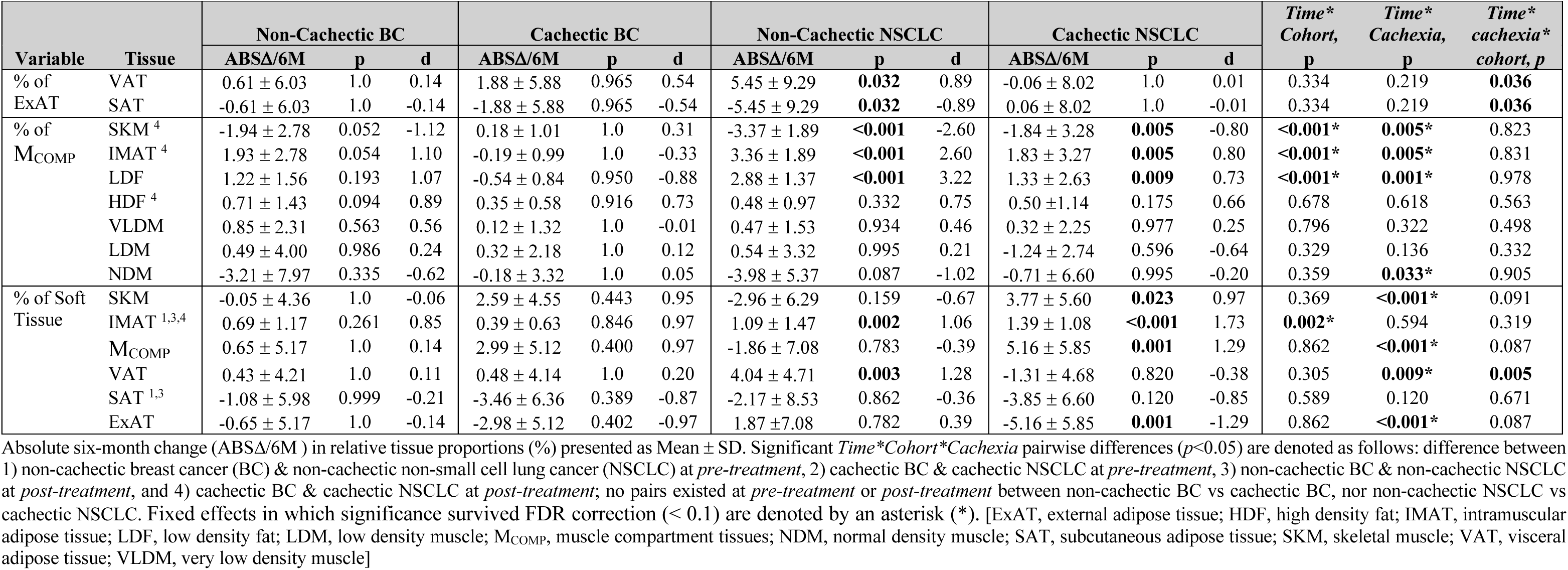
Proportional Dynamics During Cachexia Development by Cancer Type.

Numerous variables pertaining to VAT depots had a significant *Time*Cohort*Cachexia* interaction, suggesting longitudinal trends differed with cachexia development between patients with BC and NSCLC, though none survived following FDR correction. Neither cohort of BC patients demonstrated longitudinal VAT-related changes, however, divergent changes to VAT CSA and VAT CSA*HU were elucidated among NSCLC patients with the development of cachexia. Patients with NSCLC that became cachectic were characterized by declines in VAT CSA (p = 0.011), whereas those that did not develop cachexia emulated an opposing increase in CSA that neared significance (p = 0.058). When expressed as relative proportional changes across body tissues, *pre*- to *post-treatment* increases in contributions from %VAT of ExAT and soft tissue were observed only among non-cachectic patients with NSCLC, while no changes occurred among other groups.

A significant *Time*Cohort*Cachexia* interaction also existed for the M_COMP_/ExAT CSA ratio (p = 0.036), although not following FDR correction; specifically, increases in tissue quantity inside the M_COMP_, relative to quantities of adipose deposited externally, was characteristic of cachexia development only for patients with NSCLC.

Collectively, changes across all tissues combine to illustrate significant declines in body weight and BMI across both BC and NSCLC cachectic groups, as well as a significant increase among non-cachectic patients with NSCLC, each representing a large effect size, while no changes in weight or BMI were demonstrated in non-cachectic patients with BC.

## DISCUSSION

The present study provides the most detailed characterization of CT-derived body composition dynamics to date in patients with early-stage BC, with comparative evaluations against both healthy females and a cachexia-prone female cohort (NSCLC). We demonstrate herein that changes to skeletal muscle and body composition are characteristic of non-metastatic BC, which do not linearly reflect changes in body weight. Our data support that prior to initiation of NAC, non-metastatic BC patients are similar to healthy females in total muscle quantity and relative body composition. Yet, in-depth evaluations of muscle quality and composition following treatment suggest a progressive shift in tissue phenotype. This transition reflects increasing systemic physiological alterations that appear to be associated with cancer type: ranging from healthy individuals, to those with non-metastatic BC, to patients with NSCLC, a diagnosis more commonly linked with tissue wasting. Furthermore, despite *pre-treatment* differences in body and muscle composition, patients with BC and NSCLC exhibited similar morphologic profiles throughout treatment (Fig. S2). Differences between cancer types primarily concerned the magnitude of increases in IMAT tissue. While longitudinal trends were not associated with cancer type, we found distinct phenotypes corresponding with the development of cachexia, independent of cancer diagnosis. Despite presenting similarly prior to treatment, unique intra-patient patterns across adipose and muscle tissues illustrate diverging patterns of change even among patients who remain weight-stable.

While prior research has emphasized the absence of weight loss in early-stage BC,^13,37^ our data support the hypothesis that more subtle morphologic remodeling may be underway. However, these changes may only be detectable through high-resolution imaging techniques (i.e., CT), although they can be assessed across short time frames (e.g., six months). Early-stage BC patients exhibited elevations in SAT attenuation and increased intra-compartmental contributions from IMAT, despite the absence of clinically defined criteria for cachexia. The increase in SAT radiodensity may reflect localized inflammation and lipid depletion from adipocytes, both of which may correspond with adipose browning, and thus a heightened systemic metabolic demand.^38,39^

Though overlooked by conventional models for classifying cancer cachexia, early-stage BC patients experience subtle yet consistent declines in muscle quality, especially among non-cachectic patients; decreases in lean NDM and concurrent increases in lower-quality tissue within the muscle compartment suggest ongoing declines in muscle quality despite stable overall quantities. Comparatively, a decline in total compartmental quantity is characteristic of cachexia development. A shift between phenotypes of muscular alterations is illustrated by BC patients in pre-cachectic stages (Fig. 2), as proposed by Blum and Stene et al (2014) in the four-group model of cachexia.^40^ In particular, the dissociation between weight loss and the manner of muscle-specific alterations in early-stage BC aligns with a growing body of preclinical and clinical evidence that fatigue and metabolic dysfunction may precede or occur independently of substantial muscle wasting.^41^

Although total muscle quantity (e.g., M_COMP_ CSA) was comparable in healthy controls and patients with BC, their muscle quality, assessed via attenuation and subclass content, fell between healthy and NSCLC profiles (Fig. 3). Interestingly, BC patients presented with preserved levels of higher-quality muscle tissue (NDM), while also being comparable in LDM proportion, as well as LDF, HDF, VLDM, and LDM quantity, to patients with NSCLC. Elevated quantities of moderate-quality tissues may reflect increased intramyocellular lipid deposition within muscle, though in amounts subjacent to the size of image voxels.^30^ At a molecular level, we have previously shown an increase in lipid content and a corresponding altered distribution in myotubes exposed to BC conditioned media, which transpired in tandem with indicators of metabolic and mitochondrial dysfunction.^8^

The most prominent findings of the present study are the discordance between body mass changes and underlying tissue dynamics, as well as diverging phenotypes that characterize the manner of tissue-specific change. Even in the absence of weight loss, multi-faceted indicators of muscle remodeling suggest meaningful alterations to the muscular compartment that are not otherwise captured through conventional weight-based criteria, nor by many methods of body composition assessment. Specifically, cachectic patients with both BC and NSCLC experienced significant reductions in muscle and adipose tissue quantity (CSA), as well as increased SAT attenuation. In contrast, non-cachectic patients demonstrated increased proportions of intercompartmental IMAT and decrements in muscle quality, despite quantities of muscle and externally deposited adipose remaining stable. Importantly, both cachectic and non-cachectic patients exhibit comparable declines in the highest quality subclass of muscle tissue (NDM), as well as declines in metrics that combine both quantity and quality. These findings are consistent with previous reports of treatment-related fatigue and functional decline in early-stage BC patients,^42^ and align with findings of our prior work characterizing a fatigue-prone phenotype in the absence of measurable tissue wasting.^43,44^

Prior to treatment, BC patients tended to have greater amounts of VAT tissue with lower attenuations, especially among patients who evaded the development of cachexia. Recent work has supported the utility of adipose tissue in estimating the relationship between time from diagnosis and the development of cachexia across cancer stages. Specifically, patients who had metabolically active brown adipose tissue that exceeded lean tissue demands at diagnosis had better weight stability and a lower incidence of cachexia in the subsequent year.^45^ Differences in VAT tissue were found between non-cachectic patients with BC and NSCLC, both at baseline and longitudinally, although underlying alterations to muscle tissues persisted. Albeit BC patients who developed cachexia met clinical criteria for weight loss, their tissue-level changes appeared more muted than those of their NSCLC counterparts. Changes in M_COMP_ attenuation, IMAT accumulation, and ExAT remodeling were less pronounced, and in some cases, more similar to those of non-cachectic BC patients than to those of cachectic NSCLC patients. These differences were further magnified in longitudinal models evaluating the interaction of cancer type and cachexia development.

Sample-specific sources of measurement error may also play a role in elucidating the findings of the present study from those of prior research.^19^ Specifically, the bulk of extant literature estimating body composition in BC cohorts has used BIA and dual-energy x-ray absorptiometry (DXA) techniques,^46^ likely due to convenience and the limited availability of advanced imaging. Notably, BC patients and survivors commonly experience fluid imbalances (e.g., edema)^47,48^ and may have a greater prevalence of implanted foreign objects (i.e., breast implants, chemotherapy ports) compared to other clinical samples. These factors have the potential to contribute to higher rates of measurement error derived from these methods, which are common among BC-relevant cohorts. In particular, estimations of body composition have been shown to differ across various methods of assessment, including specifically among BC patients; for example, a study by Freedman and colleagues (2004) showed that despite agreement between BIA, air displacement plethysmography (ADP), and DXA prior to treatment, intra-individual changes to fat and fat-free mass throughout chemotherapy were only identified via ADP.^12^ Similarly, such methods providing whole body estimations typically express measures of fat and lean mass as relative quantities.^49^ However, we show that proportional estimations can be biased by changes in other tissues, and often do not reflect the actual measured change in tissue quantities (Fig. 1, S1, 2, S2). Techniques sensitive to compositional changes among individual tissues are likely better alternatives for assessing patient risk.

While the present study did not quantify changes in physical function, only one of the 76 patients monitored longitudinally was determined to have functional limitations prior to treatment, as suggested by their ECOG performance status. Most clinical evaluations of function, including ECOG status, aim to quantify a patient’s ability to complete activities of daily living (ADLs).^50^ Though important among many clinical samples, function captured by such tests are typically not indicative of dysfunction when implemented in early-stage BC patients, which tend to be higher-functioning than most clinical samples. Among prior work, associations between muscle and functional performance in early-stage BC have varied across methods of body composition estimation. For example, a study utilizing BIA longitudinally found functional impairments and reports of fatigue, despite stable lean and fat mass.^51^ In contrast, CT-derived assessments have suggested functional associations with muscle quality in cancer patients,^42^ including non-metastatic BC.^42,52^

Herein, we uniquely leveraged prospectively collected CT imaging obtained during a standardized treatment timeline, which afforded a higher degree of temporal control and homogeneity than studies derived from retrospective charts reviews that comprise much of the extant literature.^53–55^ As such, this allowed for the structured longitudinal study of a highly homogenous patient sample, notably during the treatment of active disease, in lieu of evaluations at follow up when most standard of care CTs are ordered for early-stage BC patients. In addition to the use of gold-standard CT imaging for segmentation of muscle and adipose tissues, classification strategies utilized herein extend beyond traditional HU thresholds to include consideration for the entire muscle compartment, as well as the distribution of comprising tissues. Using both absolute and relative tissue metrics, our findings demonstrate a discordance among these distinct presentations of body composition (Fig. 1, S1, 2, S2); as such, caution should be taken in the interpretation of relative or proportional presentations of body composition when assessed longitudinally, which may contribute to the wide variability across the literature.

Findings underscore the need for improved models for cachexia classification that incorporate tissue-level assessments of quality and proportion, rather than relying solely on weight-based thresholds for risk stratification. Prospective studies linking longitudinal CT-derived body compositional changes with symptomatic and functional outcomes, including fatigue severity and functional decline unrelated to ADL performance, are warranted to validate the clinical relevance of tissue remodeling patterns observed. Additionally, while CT segmentation allows for the quantification of tissue quantity (as CSA) and radiodensity, it does not directly assess tissue functional capacity, composition (e.g., fibrosis or lipid droplet content), or metabolic function, limiting the ability to infer mechanistic pathways driving the observed changes. As such, future work should aim to incorporate biomarkers reflective of inflammation, mitochondrial function, or lipid metabolism, and their relationship at various clinical time points with changes to imaged tissues. Interventions targeting the preservation of muscle quality, whether through exercise, pharmacologic support, or nutritional strategies, may be particularly valuable in early-stage BC, where overt wasting is uncommon, but early signs of tissue deterioration may develop into clinically meaningful patient outcomes. Ultimately, CT-derived assessments of body composition, such as those presented herein, may add value to personalized medicine strategies, particularly those tailored to prioritize quality of life. The use of similar assessments for risk stratification and intra-patient monitoring of maladaptive responses may inform treatment plan adjustments, ranging from dose reductions to the implementation of complementary therapeutics (i.e., exercise regimens or dietary planning). However, the feasibility of such efforts warrants further research.

In conclusion, this study demonstrates that early-stage BC is not exempt from meaningful alterations to body composition, despite traditional notions that link consequential wasting to fluctuations in body weight. Rather, we consistently observed declines in muscle quality and SAT quantity throughout neoadjuvant treatment in patients with BC, despite many individuals remaining weight stable. The presence of changes in muscle composition and adipose tissue morphology that occur even in the absence of substantial weight loss supports the need for a broader conceptualization of cachexia, extending beyond just weight loss and even overt muscle mass depletion. CT-derived metrics of tissue quantity, quality, and proportion offer a sensitive means of identifying subclinical tissue remodeling, which may underlie patient symptoms and functional decline in early-stage cancer patients, including BC. Collectively, these findings highlight the value of tissue-informed phenotyping and support the integration of advanced imaging metrics (i.e., CT) into future characterizations of cachexia and oncologic monitoring.

## Acknowledgments

This research was supported by the National Institutes of Arthritis, Musculoskeletal and Skin Diseases (NIAMS) under award number R01AR079445 (Pistilli).

## Competing Interests

All authors declare no financial or non-financial competing interests.

## Data Availability Statement

The datasets analyzed in the current study are available with open-access from The Cancer Imaging Archive, [https://www.cancerimagingarchive.net/collection/acrin-flt-breast/; https://www.cancerimagingarchive.net/collection/acrin-nsclc-fdg-pet/; https://www.cancerimagingarchive.net/collection/healthy-total-body-cts/].

## Notes

### Competing Interest Statement

The authors have declared no competing interest.

### Author Declarations

The source data were openly available to the public before the initiation of the study. Data were accessed in The Cancer Imaging Archive at: https://www.cancerimagingarchive.net/collection/acrin-flt-breast/; https://www.cancerimagingarchive.net/ collection/acrin-nsclc-fdg-pet/; https://www.cancerimagingarchive.net/collection/healthy-total-body-cts/

