## Supplemental Materials for "Characterization of Body Composition Dynamics Throughout Treatment in Patients with Early-Stage Breast Cancer"

**Table S1.** Imaging Parameters

| | Healthy | BC | BC 2x | BC 2x | NSCLC | NSCLC 2x | NSCLC 2x |
|---|---|---|---|---|---|---|---|
| | | Pre-Treatment | Pre-Treatment | Post-Treatment | Pre-Treatment | Pre-Treatment | Post-Treatment |
| Total (*n*) | 16 | 56 | 38 | 38 | 47 | 38 | 38 |
| Tube Voltage (kV) | 140.0 ± 0.0 [140, 140, 140] | 124.1 ± 7.3 [120, 120, 130] | 124.0 ± 7.2 [120, 120, 130] | 123.2 ± 6.6 [120, 120, 120] | 130.4 ± 8.6 [120, 130, 140] | 130.5 ± 8.4 [120, 130, 140] | 130.0 ± 8.4 [120, 130, 140] |
| Tube Current (mAmp) | 209.2 ± 126.1 [105.5, 141, 344.5] | 152.1 ± 87.6 [82, 150, 184.8] | 165.4 ± 91.4 [104.3, 150, 210.5] | 170.9 ± 92.9 [109.3, 150, 250.3] | 148.2 ± 66.0 [108, 119, 190] | 134.7 ± 53.7 [102.5, 115, 176] | 129.5 ± 50.9 [95.5, 115, 160] |
| Exposure (mAs) | 75.06 ± 45.40 [37.8, 50.5, 124] | 1879.46 ± 3413 [24.3, 75.5, 349.5] | 1952.1 ± 3501 [22.8, 78.5, 1144.3] | 1923.2 ± 3436 [23.3, 79, 1178.8] | 996.8 ± 2507 [18.3, 70, 140.5] | 804.4 ± 2186 [17, 55, 91] | 715.9 ± 2248 [13, 42, 81] |
| *Slice Thickness (n)* | | | | | | | |
| 2.344 mm | 16 | 0 | 0 | 0 | 0 | 0 | 0 |
| 3.00 mm | 0 | 0 | 0 | 0 | 2 | 2 | 3 |
| 3.75 mm | 0 | 16 | 13 | 13 | 18 | 16 | 17 |
| 4.00 mm | 0 | 12 | 6 | 5 | 1 | 1 | 1 |
| 5.00 mm | 0 | 28 | 19 | 20 | 26 | 19 | 17 |
| *Voxel Width (n)* | | | | | | | |
| 0.781 mm | 0 | 0 | 0 | 0 | 1 | 0 | 0 |
| 0.8789 mm | 0 | 0 | 0 | 0 | 1 | 0 | 0 |
| 0.9648 mm | 1 | 0 | 0 | 0 | 0 | 0 | 0 |
| 0.9766 mm | 15 | 39 | 30 | 31 | 42 | 36 | 36 |
| 1.172 mm | 0 | 5 | 1 | 1 | 1 | 0 | 0 |
| 1.367 mm | 0 | 12 | 7 | 6 | 2 | 2 | 2 |

Data presented as Mean ± Standard Deviation [Q1, median, Q3], or sample size (*n*) for all imaging samples of healthy, breast cancer (BC), and non-small cell lung cancer (NSCLC) patients.

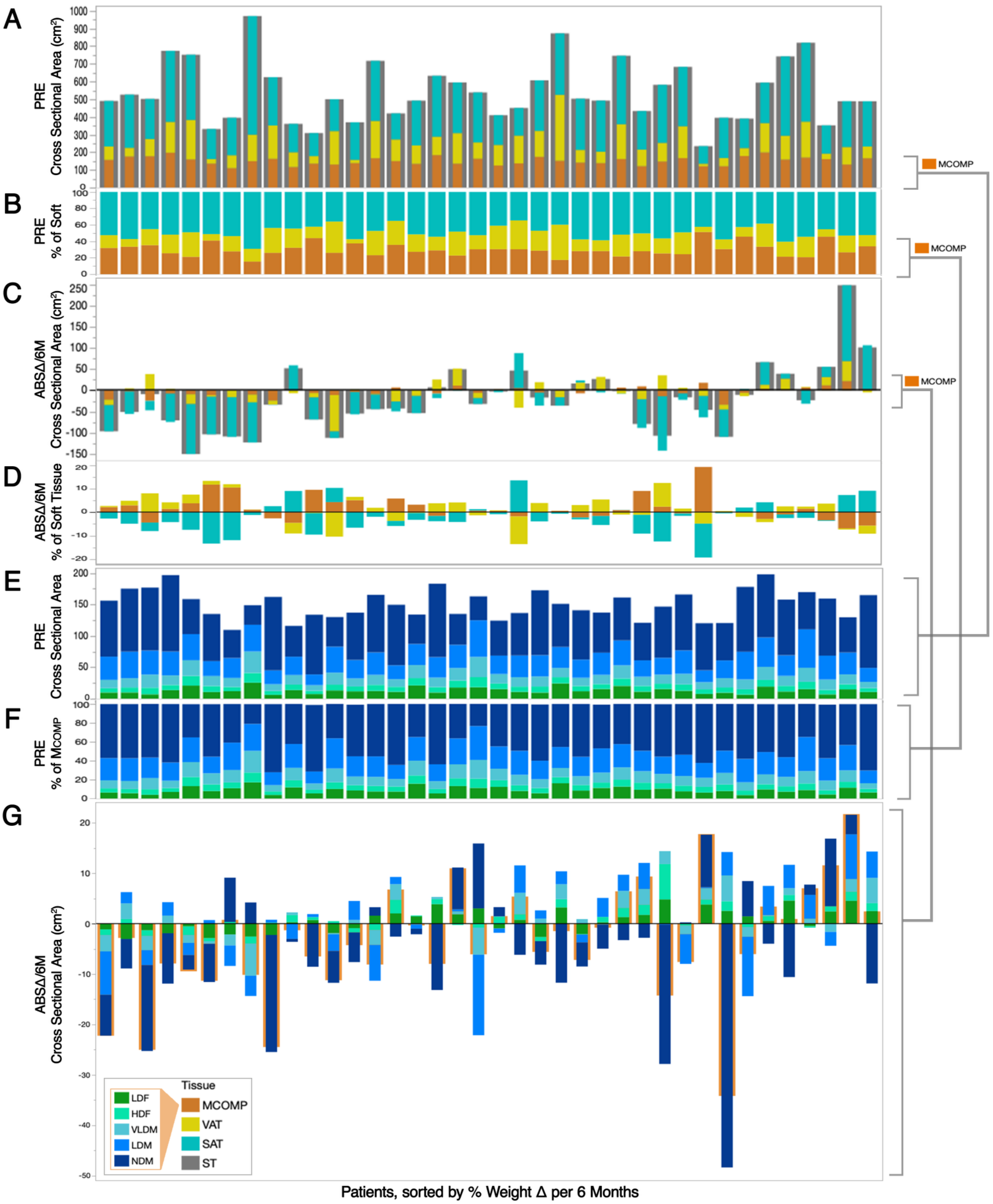


**Supplementary Figure 1A-G**. Patient Muscle Composition Profiles at Pre-Treatment and Throughout Treatment

Individual patient profiles at *pre-treatment* (A, B, E, F) and six-month changes (C, D, G) to body composition (A-D) and muscle composition (E-G) in patients with BC (n=38), in order of relative six-month change (%Δ/6M) in body weight. A) Body tissue cross-sectional area (CSA; in $cm^2$) at *pre-treatment*; B) Body tissue percentage of total soft tissue CSA (%) at *pre-treatment*; C) absolute change per six months (ABSΔ/6M) in the CSA ($cm^2$) of body tissues; D) ABSΔ/6M of body tissue proportions toward total soft tissue CSA (%); E) CSA ($cm^2$) of tissue subclasses comprising the muscle compartment ($M_{COMP}$) at *pre-treatment*; F) $M_{COMP}$ tissue subclass percentage of total $M_{COMP}$ tissue CSA (%) at *pre-treatment*; G) ABSΔ/6M in the CSA ($cm^2$) of $M_{COMP}$ tissue subclasses. [HDF, high density fat; $M_{COMP}$, muscle compartment; NDM, normal density muscle; LDF, low density fat; LDM, low density muscle; SAT, subcutaneous adipose tissue; ST, soft tissue; VAT, visceral adipose tissue; VLDM, very low density muscle]

**Table S2.** Mixed Model Fixed Effect Statistics

| Variable | ME *Time* | ME *Cohort* | ME *Cachexia* | *Cohort*Cachexia* | *Time*Cohort* | *Time*Cachexia* | *Time*Cohort *Cachexia* | Wald *p*-value | % Variance from ID | % Variance from Residual |
|---|---|---|---|---|---|---|---|---|---|---|
| Body Mass (kg) [1,3,6,7,8] | **<0.001*** | **<0.001*** | 0.111 | **0.011** | 0.095 | **<0.001*** | 0.054 | **<0.001** | 97.85 | 2.15 |
| BMI (kg/m$^2$) [1,3,6,7,8] | **<0.001*** | **<0.001*** | 0.220 | **0.009** | 0.108 | **<0.001*** | **0.036** | **<0.001** | 97.81 | 2.19 |
| SKM CSA [1,3,6,8] | **<0.001*** | **<0.001*** | 0.947 | 0.471 | 0.710 | **0.002*** | 0.985 | **<0.001** | 94.82 | 5.18 |
| IMAT CSA [7] | **<0.001*** | 0.733 | 0.651 | 0.107 | **0.002*** | **<0.001*** | 0.722 | **<0.001** | 92.45 | 7.55 |
| $M_{COMP}$ CSA [1,3,6,8] | **0.003*** | **<0.001*** | 0.910 | 0.194 | 0.237 | **<0.001*** | 0.876 | **<0.001** | 94.73 | 5.27 |
| VAT CSA [8] | 0.497 | 0.100 | 0.807 | 0.226 | 0.647 | **<0.001*** | **0.028** | **<0.001** | 93.14 | 6.86 |
| SAT CSA [1,3,6,8] | **<0.001*** | **<0.001*** | 0.271 | 0.075 | 0.745 | **<0.001*** | 0.922 | **<0.001** | 91.90 | 8.10 |
| ExAT CSA [1,3,6,8] | **0.001*** | **0.001*** | 0.511 | 0.078 | 0.652 | **<0.001*** | 0.358 | **<0.001** | 93.27 | 6.73 |
| Soft Tissue CSA [1,3,6,8] | **<0.001*** | **<0.001*** | 0.532 | 0.071 | 0.560 | **<0.001*** | 0.381 | **<0.001** | 93.86 | 6.14 |
| LDF CSA [7] | **<0.001*** | 0.982 | 0.636 | 0.110 | **<0.001*** | **<0.001*** | 0.790 | **<0.001** | 92.11 | 7.89 |
| HDF CSA | **0.009*** | 0.280 | 0.733 | 0.158 | 0.564 | **0.023*** | 0.673 | **<0.001** | 90.00 | 10.00 |
| VLDM CSA | 0.404 | 0.810 | 0.383 | 0.262 | 0.481 | **0.008*** | 0.661 | **<0.001** | 90.34 | 9.66 |
| LDM CSA [8] | 0.288 | 0.103 | 0.824 | 0.094 | 0.653 | **0.002*** | 0.280 | **<0.001** | 90.00 | 10.00 |
| NDM CSA [2] | **<0.001*** | **<0.001*** | 0.703 | 0.809 | 0.868 | 0.705 | 0.756 | **<0.001** | 91.64 | 8.36 |
| NDM mean HU | 0.228 | **<0.001*** | 0.541 | 0.563 | 0.334 | 0.889 | 0.476 | **<0.001** | 74.68 | 25.32 |
| SKM mean HU | **0.009*** | **0.012*** | 0.537 | 0.344 | 0.699 | 0.193 | 0.779 | **<0.001** | 87.14 | 12.86 |
| $M_{COMP}$ mean HU [4,7] | **<0.001*** | **0.020*** | 0.583 | 0.250 | **0.029** | **0.015*** | 0.899 | **<0.001** | 90.58 | 9.42 |
| VAT mean HU | 0.272 | **0.027*** | 0.764 | 0.437 | 0.112 | **0.001*** | 0.100 | **<0.001** | 85.12 | 14.88 |
| SAT mean HU [6,8] | **<0.001*** | **0.024*** | 0.689 | 0.815 | 0.192 | **<0.001*** | 0.395 | **<0.001** | 82.11 | 17.89 |
| ExAT mean HU [6,8] | **<0.001*** | **0.007*** | 0.745 | 0.956 | 0.150 | **<0.001*** | 0.350 | **<0.001** | 82.98 | 17.02 |
| NDM CSA*HU [1,3] | **<0.001*** | **<0.001*** | 0.819 | 0.728 | 0.754 | 0.731 | 0.898 | **<0.001** | 90.57 | 9.43 |
| SKM CSA*HU [2,3,4] | **<0.001*** | **<0.001*** | 0.756 | 0.787 | 0.906 | 0.645 | 0.938 | **<0.001** | 91.54 | 8.46 |
| $M_{COMP}$ CSA*HU [4,7] | **<0.001*** | **0.001*** | 0.647 | 0.397 | 0.131 | 0.245 | 0.952 | **<0.001** | 91.89 | 8.11 |
| VAT CSA*HU [7,8] | 0.469 | 0.094 | 0.737 | 0.243 | 0.498 | **<0.001*** | **0.034** | **<0.001** | 93.24 | 6.76 |
| SAT CSA*HU [1,3,6,8] | **<0.001*** | **<0.001*** | 0.242 | 0.133 | 0.465 | **<0.001*** | 0.683 | **<0.001** | 91.42 | 8.58 |
| SKM SMI (cm$^2$/m$^2$) [1,3,6,8] | **<0.001*** | **<0.001*** | 0.662 | 0.464 | 0.691 | **0.003*** | 0.949 | **<0.001** | 94.61 | 5.39 |
| SKM SMG | **<0.001*** | **<0.001*** | 0.616 | 0.759 | 0.889 | 0.693 | 0.951 | **<0.001** | 91.18 | 8.82 |
| %SAT of ExAT [7] | **0.016*** | 0.103 | 0.192 | 0.638 | 0.334 | 0.219 | **0.036** | **<0.001** | 85.36 | 14.64 |
| %SKM of $M_{COMP}$ [4,7,8] | **<0.001*** | **0.042*** | 0.698 | 0.305 | **<0.001*** | **0.005*** | 0.823 | **<0.001** | 92.56 | 7.44 |
| %LDF of $M_{COMP}$ [7,8] | **<0.001*** | 0.235 | 0.660 | 0.269 | **<0.001*** | **0.001*** | 0.978 | **<0.001** | 92.13 | 7.87 |
| %HDF of $M_{COMP}$ [4] | **<0.001*** | **0.001*** | 0.833 | 0.501 | 0.678 | 0.618 | 0.563 | **<0.001** | 88.78 | 11.22 |
| %VLDM of $M_{COMP}$ | 0.065* | **0.009*** | 0.368 | 0.676 | 0.796 | 0.322 | 0.498 | **<0.001** | 86.93 | 13.07 |
| %LDM of $M_{COMP}$ | 0.967 | 0.252 | 0.631 | 0.272 | 0.329 | 0.136 | 0.332 | **<0.001** | 84.17 | 15.83 |
| %NDM of $M_{COMP}$ | **0.007*** | **0.018*** | 0.518 | 0.294 | 0.359 | **0.033*** | 0.905 | **<0.001** | 88.84 | 11.16 |
| %SKM of Soft Tissue [8] | 0.183 | **0.043*** | 0.568 | 0.159 | 0.369 | **<0.001*** | 0.091 | **<0.001** | 91.58 | 8.42 |
| %IMAT of Soft Tissue [1,3,4,7,8] | **<0.001*** | **<0.001*** | 0.539 | 0.517 | **0.002*** | 0.594 | 0.319 | **<0.001** | 88.33 | 11.67 |
| %VAT of Soft Tissue [7] | 0.076* | 0.782 | 0.421 | 0.842 | 0.305 | **0.009*** | **0.005** | **<0.001** | 91.38 | 8.62 |
| %SAT of Soft Tissue [1,3] | **0.001*** | **0.001*** | 0.130 | 0.131 | 0.589 | 0.120 | 0.671 | **<0.001** | 83.14 | 16.86 |
| % $M_{COMP}$ of Soft Tissue [8] | **0.012*** | **0.010*** | 0.528 | 0.159 | 0.862 | **<0.001*** | 0.087 | **<0.001** | 91.16 | 8.84 |
| VAT/SAT CSA ratio [3] | 0.082* | 0.101 | 0.174 | 0.430 | 0.244 | 0.242 | 0.087 | **<0.001** | 77.99 | 22.01 |
| SKM/IMAT CSA ratio [4,5,7] | **<0.001*** | **0.011*** | 0.902 | 0.146 | **0.011*** | **<0.001*** | 0.551 | **<0.001** | 90.95 | 9.05 |
| $M_{COMP}$/ExAT CSA ratio [8] | 0.050* | **0.018*** | 0.749 | 0.398 | 0.945 | **0.001*** | **0.036** | **<0.001** | 88.23 | 11.77 |

| | | | | | | | | | | |
|---|---|---|---|---|---|---|---|---|---|---|
| SKM-$M_{COMP}$ HU difference [7,8] | **<0.001*** | 0.151 | 0.768 | 0.264 | **<0.001*** | **0.001*** | 0.834 | **<0.001** | 91.64 | 8.36 |
| VAT-SAT HU difference | **0.001*** | 0.856 | 0.304 | 0.130 | 0.753 | 0.476 | 0.351 | **<0.001** | 75.03 | 24.97 |

P-values for all mixed model fixed effects, as well as random effect statistics regarding patient-sourced variance (Variance from ID, %) and other sources of variance (%). Additionally, significant *Time*Cohort*Cachexia* pairwise differences are denoted in superscript as follows: 1) breast cancer (BC) non-cachectic vs non-small cell lung cancer (NSCLC) non-cachectic @ pre, 2) BC cachectic vs NSCLC cachectic @ pre, 3) BC non-cachectic vs NSCLC non-cachectic @ post, 4) BC cachectic vs NSCLC cachectic @ post, 5) BC non-cachectic pre vs post, 6) BC cachectic pre vs post, 7) NSCLC non-cachectic pre vs post, 8) NSCLC cachectic pre vs post; no paired differences existed at *pre-treatment* or *post-treatment* for NSCLC non-cachectic vs NSCLC cachectic or BC non-cachectic vs BC cachectic. Fixed effects in which significance survived FDR correction (< 0.1) are denoted by an asterisk (*). [BMI, body mass index; CSA, cross-sectional surface area; CSA*HU, product of cross-sectional area and mean attenuation; ExAT, external adipose tissue; HDF, high density fat; HU, Hounsfield units; IMAT, intramuscular adipose tissue; LBM, lean body mass; LDF, low density fat; LDM, low density muscle; $M_{COMP}$, muscle compartment tissues; NDM, normal density muscle; SAT, subcutaneous adipose tissue; SKM, skeletal muscle; SMG, skeletal muscle gauge; SMI, skeletal muscle index; VAT, visceral adipose tissue; VLDM, very low density muscle]

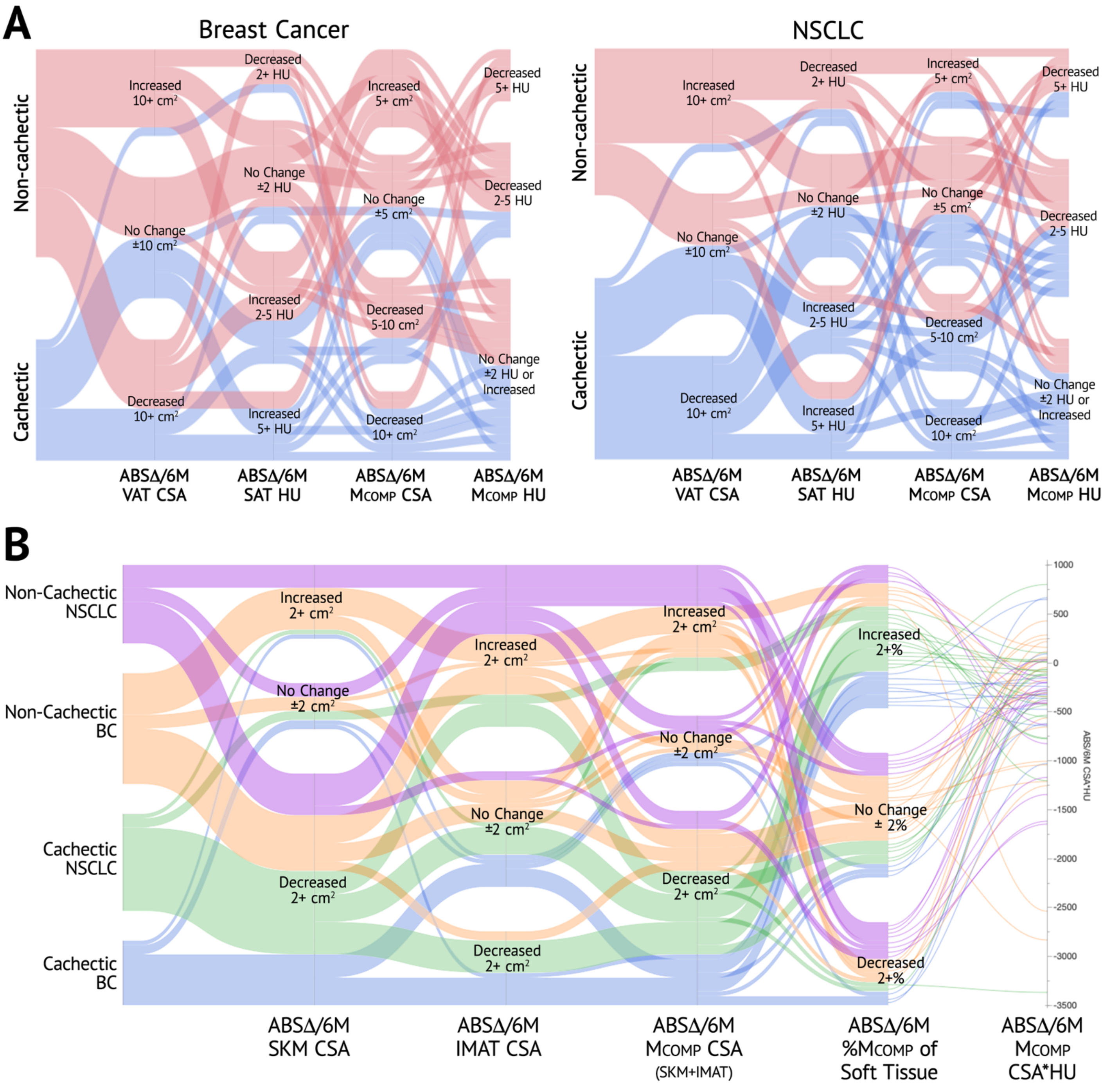


**Supplementary Figure S2**. Changes to Body and Muscle Tissues in Patients with Breast Cancer and NSCLC

Summary of cachectic and non-cachectic trends in A) body composition changes, and B) muscle compartment ($M_{COMP}$) changes, in patients with BC and NSCLC from *pre-* to *post-treatment*. [ABSΔ/6M, absolute change per six months; CSA, cross-sectional area; CSA*HU, product of tissue cross-sectional area and mean attenuation; HU, Hounsfield units; IMAT, intramuscular adipose tissue; $M_{COMP}$, muscle compartment; SAT, subcutaneous adipose tissue; VAT, visceral adipose tissue]

**Table S3.** Tissue Changes Throughout Non-Surgical Treatment in Patients with BC and NSCLC

| | BC | | | | NSCLC | | | | ME | ME | *Time** |
|---|---|---|---|---|---|---|---|---|---|---|---|
| **Tissue Variable** | **Pre-Treatment** | **ABSΔ/6M** | **p** | **d** | **Pre-Treatment** | **ABSΔ/6M** | **p** | **d** | ***Time, p*** | ***Cohort, p*** | ***Cohort, p*** |
| Weight (kg) [1,2] | 79.83 ± 13.81 | -2.90 ± 4.63 | **<0.001** | -0.89 | 65.96 ± 14.08 | -2.97 ± 5.98 | **<0.001** | -0.70 | **<0.001*** | **<0.001*** | 0.095 |
| BMI (kg/m$^2$) [1,2] | 30.33 ± 5.48 | -1.11 ± 1.75 | **<0.001** | -0.90 | 25.66 ± 5.20 | -1.16 ± 2.34 | **<0.001** | -0.70 | **<0.001*** | **<0.001*** | 0.108 |
| SKM CSA (cm$^2$) [1,2] | 128.4 ± 23.06 | -4.63 ± 10.85 | **0.004** | -0.60 | 105.1 ± 16.09 | -5.11 ± 6.70 | **<0.001** | -1.08 | **<0.001*** | **<0.001*** | 0.710 |
| IMAT CSA (cm$^2$) | 22.40 ± 6.86 | 1.18 ± 3.41 | 0.944 | 0.49 | 23.97 ± 9.71 | 2.95 ± 4.82 | **<0.001** | 0.87 | **<0.001*** | 0.733 | **0.002*** |
| VAT CSA (cm$^2$) | 119.0 ± 73.06 | -0.85 ± 24.91 | 0.856 | -0.05 | 95.54 ± 62.12 | -2.62 ± 36.53 | 0.999 | -0.10 | 0.497 | 0.100 | 0.647 |
| SAT CSA (cm$^2$) [1,2] | 271.4 ± 109.9 | -19.48 ± 60.02 | **0.011** | -0.46 | 186.9 ± 107.0 | -25.02 ± 53.20 | **0.032** | -0.67 | **<0.001*** | **<0.001*** | 0.745 |
| $M_{COMP}$ CSA (cm$^2$) [1,2] | 150.8 ± 22.39 | -3.43 ± 11.49 | **0.020** | -0.42 | 129.1 ± 19.95 | -2.15 ± 8.44 | 0.529 | -0.36 | **0.003*** | **<0.001*** | 0.237 |
| ExAT CSA (cm$^2$) [1,2] | 390.4 ± 158.7 | -20.33 ± 69.55 | **0.035** | -0.41 | 282.5 ± 150.3 | -27.64 ± 78.40 | 0.132 | -0.50 | **<0.001*** | **0.001*** | 0.652 |
| Soft tissue CSA (cm$^2$) [1,2] | 541.2 ± 168.9 | -23.77 ± 75.27 | **0.016** | -0.45 | 411.6 ± 161.9 | -29.80 ± 81.90 | 0.105 | -0.51 | **<0.001*** | **<0.001*** | 0.560 |
| LDF CSA (cm$^2$) | 13.17 ± 4.89 | 0.60 ± 2.30 | 1.000 | 0.37 | 13.76 ± 6.76 | 2.47 ± 3.63 | **<0.001** | 0.96 | **<0.001*** | 0.982 | **<0.001*** |
| HDF CSA (cm$^2$) | 9.23 ± 2.44 | 0.58 ± 1.55 | 0.470 | 0.53 | 10.21 ± 3.26 | 0.47 ± 1.61 | 0.100 | 0.42 | **0.009*** | 0.280 | 0.564 |
| VLDM CSA (cm$^2$) | 16.68 ± 5.23 | 0.39 ± 2.61 | 0.999 | 0.21 | 17.51 ± 5.35 | 0.32 ± 2.69 | 0.684 | 0.17 | 0.404 | 0.810 | 0.481 |
| LDM CSA (cm$^2$) | 34.42 ± 9.37 | -0.19 ± 5.08 | 0.973 | -0.05 | 31.79 ± 7.60 | -1.03 ± 4.47 | 0.695 | -0.32 | 0.288 | 0.104 | 0.653 |
| NDM CSA (cm$^2$) [1,2] | 76.99 ± 24.36 | -4.70 ± 11.51 | 0.058 | -0.58 | 55.17 ± 15.21 | -4.05 ± 8.52 | **0.026** | -0.67 | **<0.001*** | **<0.001*** | 0.868 |
| NDM mean HU [1,2] | 49.58 ± 3.26 | -0.72 ± 2.70 | 0.429 | -0.38 | 47.20 ± 2.68 | -0.06 ± 2.34 | 0.998 | -0.03 | 0.228 | **<0.001*** | 0.334 |
| SKM mean HU [2] | 32.48 ± 7.01 | -1.39 ± 4.11 | 0.378 | -0.48 | 28.12 ± 5.81 | -1.14 ± 3.65 | 0.128 | -0.44 | **0.009*** | **0.013*** | 0.699 |
| $M_{COMP}$ mean HU [2] | 18.80 ± 10.11 | -2.12 ± 5.10 | 0.361 | -0.59 | 12.51 ± 8.89 | -3.34 ± 5.11 | **<0.001** | -0.92 | **<0.001*** | **0.020*** | **0.029** |
| VAT mean HU | -95.37 ± 6.71 | 1.39 ± 5.88 | 0.240 | 0.33 | -91.63 ± 8.85 | 0.11 ± 6.29 | 0.984 | 0.03 | 0.272 | **0.027*** | 0.112 |
| SAT mean HU | -105.2 ± 5.43 | 2.46 ± 4.72 | **0.003** | 0.74 | -101.9 ± 8.80 | 1.88 ± 6.66 | 0.266 | 0.40 | **<0.001*** | **0.024*** | 0.192 |
| ExAT mean HU [1] | -102.7 ± 5.12 | 2.34 ± 4.47 | **0.003** | 0.74 | -98.72 ± 8.48 | 1.60 ± 6.31 | 0.369 | 0.36 | **<0.001*** | **0.007*** | 0.150 |
| NDM CSA*HU [1,2] | 3863 ± 1367 | -300.0 ± 648.0 | **0.030** | -0.65 | 2627 ± 819.7 | -201.4 ± 508.7 | 0.073 | -0.56 | **<0.001*** | **<0.001*** | 0.754 |
| SKM CSA*HU [1,2] | 4239 ± 1397 | -321.8 ± 644.9 | **0.014** | -0.71 | 2976 ± 860.1 | -272.3 ± 495.4 | **0.007** | -0.78 | **<0.001*** | **<0.001*** | 0.906 |
| $M_{COMP}$ CSA*HU [1,2] | 2903 ± 1726 | -378.5 ± 721.5 | 0.078 | -0.74 | 1575 ± 1241 | -477.4 ± 689.9 | **<0.001** | -0.98 | **<0.001*** | **<0.001*** | 0.131 |
| VAT CSA*HU | -11651 ± 7627 | 160.3 ± 2534 | 0.760 | 0.09 | -9133 ± 6278 | 208.0 ± 3789 | 1.000 | 0.08 | 0.469 | 0.094 | 0.498 |
| SAT CSA*HU [1,2] | -28800 ± 12960 | 2583 ± 6901 | **0.002** | 0.53 | -19569 ± 11973 | 2877 ± 5997 | **0.034** | 0.68 | **<0.001*** | **<0.001*** | 0.465 |
| SMI (cm$^2$/m$^2$) [1,2] | 48.75 ± 8.78 | -1.79 ± 4.20 | **0.005** | -0.60 | 41.02 ± 6.54 | -1.99 ± 2.64 | **<0.001** | -1.07 | **<0.001*** | **<0.001*** | 0.691 |
| SMG [1,2] | 1611 ± 543.4 | -124.8 ± 252.6 | **0.017** | -0.70 | 1160 ± 333.1 | -105.8 ± 196.0 | **0.007** | -0.76 | **<0.001*** | **<0.001*** | 0.889 |
| VAT/SAT CSA ratio | 0.447 ± 0.253 | 0.012 ± 0.151 | 0.976 | 0.11 | 0.552 ± 0.303 | 0.064 ± 0.277 | 0.159 | 0.33 | 0.082* | 0.101 | 0.244 |
| SKM/IMAT CSA ratio [2] | 6.31 ± 2.37 | -0.470 ± 1.052 | 0.364 | -0.63 | 4.91 ± 1.66 | -0.769 ± 1.250 | **<0.001** | -0.87 | **<0.001*** | **0.011*** | **0.011*** |
| $M_{COMP}$/ExAT CSA ratio | 0.447 ± 0.187 | 0.055 ± 0.191 | 0.541 | 0.41 | 0.653 ± 0.477 | 0.081 ± 0.356 | 0.454 | 0.32 | 0.050* | **0.018*** | 0.945 |
| SKM-$M_{COMP}$ HU difference [2] | 13.68 ± 4.04 | 0.73 ± 1.91 | 0.815 | 0.54 | 15.60 ± 4.42 | 2.20 ± 2.58 | **<0.001** | 1.21 | **<0.001*** | 0.151 | **<0.001*** |
| VAT-SAT HU difference | 9.82 ± 5.88 | 1.08 ± 5.23 | 0.157 | 0.29 | 10.32 ± 4.85 | 1.77 ± 4.05 | **0.049** | 0.62 | **0.001*** | 0.856 | 0.753 |

Pre-treatment and absolute six-month change (ABSΔ/6M) in cross-sectional surface area (cm$^2$), mean attenuation (HU), tissue quantity-quality products (cm$^2$*HU), and other tissue parameters presented as Mean ± SD. Significant *Time*Cohort* paired differences ($p$<0.05) are denoted as follows: difference between breast cancer (BC) & non-small cell lung cancer (NSCLC) at 1) *pre-treatment*, and 2) *post-treatment*. Fixed effects in which significance survived FDR correction (< 0.1) are denoted by an asterisk (*). [BMI, body mass index; CSA, cross-sectional surface area; ExAT, external adipose tissue; HDF, high density fat; HU, Hounsfield units; IMAT, intramuscular adipose tissue; LBM, lean body mass; LDF, low density fat; LDM, low density muscle; $M_{COMP}$, muscle compartment tissues; ME, main effect; NDM, normal density muscle; SAT, subcutaneous adipose tissue; SKM, skeletal muscle; SMG, skeletal muscle gauge; SMI, skeletal muscle index; VAT, visceral adipose tissue; VLDM, very low density muscle]

**Table S4.** Proportional Tissue Changes Throughout Non-Surgical Treatment in Patients with BC and NSCLC

| Parent | Tissue | BC | | | | NSCLC | | | | ME | ME | *Time*Cohort* |
|---|---|---|---|---|---|---|---|---|---|---|---|---|
| | | **Pre-Treatment** | **ABSΔ/6M** | **p** | **d** | **Pre-Treatment** | **ABSΔ/6M** | **p** | **d** | **Time, *p*** | **Cohort, *p*** | **IxD, *p*** |
| % of ExAT | VAT | 28.99 ± 11.19 | 1.08 ± 5.93 | 0.729 | 0.26 | 33.45 ± 11.62 | 2.41 ± 8.93 | 0.075 | 0.38 | **0.016*** | 0.103 | 0.334 |
| | SAT | 71.01 ± 11.19 | -1.08 ± 5.93 | 0.729 | -0.26 | 66.55 ± 11.62 | -2.41 ± 8.93 | 0.075 | -0.38 | **0.016*** | 0.103 | 0.334 |
| % of $M_{COMP}$ | SKM [2] | 84.88 ± 4.84 | -1.15 ± 2.49 | 0.375 | -0.66 | 81.67 ± 5.64 | -2.52 ± 2.82 | **<0.001** | -1.26 | **<0.001*** | **0.042*** | **<0.001*** |
| | IMAT [2] | 15.13 ± 4.84 | 1.15 ± 2.49 | 0.383 | 0.65 | 18.33 ± 5.65 | 2.51 ± 2.82 | **<0.001** | 1.26 | **<0.001*** | **0.042*** | **<0.001*** |
| | LDF [2] | 8.93 ± 3.46 | 0.57 ± 1.58 | 0.914 | 0.51 | 10.46 ± 4.14 | 2.02 ± 2.27 | **<0.001** | 1.26 | **<0.001*** | 0.235 | **<0.001*** |
| | HDF [1,2] | 6.20 ± 1.64 | 0.57 ± 1.19 | 0.039 | 0.69 | 7.87 ± 1.85 | 0.49 ± 1.05 | **0.006** | 0.66 | **<0.001*** | **<0.001*** | 0.679 |
| | VLDM [1,2] | 11.19 ± 3.45 | 0.58 ± 2.02 | 0.676 | 0.41 | 13.53 ± 3.14 | 0.39 ± 1.94 | 0.425 | 0.28 | 0.065 | **0.009*** | 0.796 |
| | LDM | 22.90 ± 5.41 | 0.43 ± 3.41 | 0.891 | 0.18 | 24.58 ± 4.18 | -0.45 ± 3.10 | 0.906 | -0.20 | 0.967 | 0.252 | 0.329 |
| | NDM [2] | 50.58 ± 12.13 | -2.09 ± 6.75 | 0.567 | -0.44 | 43.05 ± 10.18 | -2.17 ± 6.22 | **0.046** | -0.49 | **0.007*** | **0.018*** | 0.359 |
| % of Soft Tissue | SKM | 25.59 ± 7.72 | 0.93 ± 4.56 | 0.405 | 0.29 | 29.59 ± 12.14 | 0.76 ± 6.75 | 0.989 | 0.16 | 0.183 | **0.043*** | 0.369 |
| | IMAT [1,2] | 4.30 ± 1.12 | 0.58 ± 1.01 | 0.057 | 0.81 | 6.29 ± 2.29 | 1.26 ± 1.26 | **<0.001** | 1.41 | **<0.001*** | **<0.001*** | **0.002*** |
| | $M_{COMP}$ [1] | 29.89 ± 7.96 | 1.51 ± 5.21 | 0.229 | 0.41 | 35.88 ± 13.35 | 2.02 ± 7.26 | 0.316 | 0.39 | **0.012*** | **0.010*** | 0.862 |
| | VAT | 20.70 ± 8.93 | 0.45 ± 4.13 | 0.951 | 0.15 | 21.59 ± 8.98 | 1.08 ± 5.36 | 0.185 | 0.29 | 0.076 | 0.782 | 0.305 |
| | SAT [1,2] | 49.41 ± 7.67 | -1.95 ± 6.15 | 0.185 | -0.45 | 42.52 ± 11.10 | -3.10 ± 7.46 | **0.026** | -0.59 | **<0.001*** | **<0.001*** | 0.589 |
| | ExAT [1] | 70.11 ± 7.97 | -1.51 ± 5.21 | 0.230 | -0.41 | 64.11 ± 13.35 | -2.02 ± 7.26 | 0.318 | -0.39 | **0.012*** | **0.010*** | 0.862 |

Pre-treatment and absolute six-month change (ABSΔ/6M) in relative tissue proportions (%) presented as Mean ± SD. Significant *Time*Cohort* paired differences ($p$<0.05) are denoted as follows: difference between breast cancer (BC) & non-small cell lung cancer (NSCLC) at 1) *pre-treatment*, and 2) *post-treatment*. Fixed effects in which significance survived FDR correction (< 0.1) are denoted by an asterisk (*). [ExAT, external adipose tissue; HDF, high density fat; IMAT, intramuscular adipose tissue; IxD, interaction; LDF, low density fat; LDM, low density muscle; $M_{COMP}$, muscle compartment tissues; ME, main effect; NDM, normal density muscle; SAT, subcutaneous adipose tissue; SKM, skeletal muscle; VAT, visceral adipose tissue; VLDM, very low density muscle]

**Table S5.** Proportional Changes Throughout Non-Surgical Treatment in Cachectic and Non-Cachectic Patients

| | | Non-Cachectic | | | | Cachectic | | | | ME | ME | *Time*Cachexia* |
|---|---|---|---|---|---|---|---|---|---|---|---|---|
| | **Tissue** | **Pre-Treatment** | **ABSΔ/6M** | **p** | **d** | **Pre-Treatment** | **ABSΔ/6M** | **p** | **d** | ***Time*** | ***Cachexia*** | **IxD, *p*** |
| % of ExAT | VAT | 32.23 ± 11.83 | 2.62 ± 7.83 | **0.038** | 0.47 | 30.04 ± 11.27 | 0.72 ± 7.21 | 0.841 | 0.14 | **0.016*** | 0.192 | 0.219 |
| | SAT | 67.77 ± 11.83 | -2.62 ± 7.83 | **0.038** | -0.47 | 69.96 ± 11.27 | -0.72 ± 7.21 | 0.841 | -0.14 | **0.016*** | 0.192 | 0.219 |
| % of $M_{COMP}$ | SKM | 83.92 ± 4.74 | -2.53 ± 2.53 | **<0.001** | -1.42 | 82.52 ± 6.20 | -1.03 ± 2.78 | 0.133 | -0.52 | **<0.001*** | 0.698 | **0.005*** |
| | IMAT | 16.08 ± 4.74 | 2.52 ± 2.52 | **<0.001** | 1.41 | 17.49 ± 6.20 | 1.02 ± 2.77 | 0.138 | 0.52 | **<0.001*** | 0.694 | **0.005*** |
| | LDF | 9.27 ± 3.50 | 1.91 ± 1.68 | **<0.001** | 1.60 | 10.19 ± 4.25 | 0.58 ± 2.28 | 0.449 | 0.36 | **<0.001*** | 0.660 | **0.001*** |
| | HDF | 6.81 ± 1.63 | 0.61 ± 1.25 | **0.003** | 0.70 | 7.30 ± 2.22 | 0.44 ± 0.94 | 0.053 | 0.66 | **<0.001*** | 0.833 | 0.618 |
| | VLDM | 11.80 ± 3.03 | 0.69 ± 2.01 | 0.156 | 0.49 | 13.01 ± 3.90 | 0.24 ± 1.91 | 0.933 | 0.18 | 0.065* | 0.368 | 0.322 |
| | LDM | 23.25 ± 5.24 | 0.51 ± 3.69 | 0.663 | 0.19 | 24.31 ± 4.42 | -0.62 ± 2.61 | 0.753 | -0.33 | 0.967 | 0.631 | 0.136 |
| | NDM | 48.51 ± 10.96 | -3.53 ± 6.94 | **0.003** | -0.72 | 44.83 ± 12.48 | -0.50 ± 5.47 | 0.976 | -0.13 | **0.007*** | 0.518 | **0.033*** |
| % of Soft Tissue | SKM | 28.14 ± 10.11 | -1.25 ± 5.37 | 0.162 | -0.33 | 26.94 ± 10.64 | 3.30 ± 5.17 | **0.002** | 0.90 | 0.183 | 0.568 | **<0.001*** |
| | IMAT | 5.25 ± 2.22 | 0.86 ± 1.30 | **<0.001** | 0.93 | 5.36 ± 1.86 | 0.99 ± 1.04 | **<0.001** | 1.34 | **<0.001*** | 0.539 | 0.594 |
| | $M_{COMP}$ | 33.39 ± 11.27 | -0.39 ± 6.08 | 0.780 | -0.09 | 32.29 ± 11.53 | 4.29 ± 5.60 | **<0.001** | 1.08 | **0.012*** | 0.528 | **<0.001*** |
| | VAT | 21.66 ± 8.92 | 1.92 ± 4.72 | **0.008** | 0.58 | 20.55 ± 8.98 | -0.60 ± 4.49 | 0.932 | -0.19 | 0.076* | 0.421 | **0.009*** |
| | SAT | 44.95 ± 9.77 | -1.53 ± 7.07 | 0.505 | -0.31 | 47.16 ± 10.46 | -3.69 ± 6.41 | **0.006** | -0.81 | **0.001*** | 0.130 | 0.120 |
| | ExAT | 66.61 ± 11.27 | 0.40 ± 6.08 | 0.779 | 0.09 | 67.71 ± 11.53 | -4.29 ± 5.60 | **<0.001** | -1.08 | **0.012*** | 0.528 | **<0.001*** |

Pre-treatment and absolute six-month change (ABSΔ/6M) in relative tissue proportions (%) presented as Mean ± SD, as determined using Slice-O-Matic. No significant *Timepoint*Cachexia* differences ($p$<0.05) existed at *pre-treatment* or *post-treatment* between non-cachectic and cachectic patients. Fixed effects in which significance survived FDR correction (< 0.1) are denoted by an asterisk (*). [ExAT, external adipose tissue; HDF, high density fat; IMAT, intramuscular adipose tissue; LDF, low density fat; LDM, low density muscle; $M_{COMP}$, muscle compartment tissues; NDM, normal density muscle; SAT, subcutaneous adipose tissue; SKM, skeletal muscle; VAT, visceral adipose tissue; VLDM, very low density muscle]
